# Harmonic patterns embedded in ictal EEG signals in focal epilepsy: new insight into the epileptogenic zone

**DOI:** 10.1101/2023.12.20.23300274

**Authors:** Lingli Hu, Lingqi Ye, Hongyi Ye, Xiaochen Liu, Kai Xiong, Yuanming Zhang, Zhe Zheng, Hongjie Jiang, Cong Chen, Zhongjin Wang, Jiping Zhou, Yingcai Wu, Kejie Huang, Junming Zhu, Zhong Chen, Meiping Ding, Dongping Yang, Shuang Wang

## Abstract

**Objective:** Localization of the epileptogenic zone (EZ) requires further refinement. We identified a unique ictal spectral structure, the ‘harmonic pattern’ (H pattern), which potentially serves as a novel biomarker for localizing the EZ. This study aimed to analyze the clinical significance of the H pattern and to explore its underlying waveform features.

**Methods:** Seventy patients with drug-resistant focal epilepsy, undergoing stereo-EEG (SEEG) evaluation and surgery, were included. Time-frequency maps (TFM) were generated using Morlet wavelet transform analysis. The H pattern was defined as multiple equidistant, high-density bands with varying frequencies on TFM. The upper quartile was employed to confirm contacts expressing dominant H pattern (*d*H pattern). Bispectral analysis and transfer function modeling were employed to assess nonlinear properties and propagation, respectively. The performance of the *d*H pattern in evaluating the EZ was compared with other ictal biomarkers.

**Results:** Regardless of seizure onset patterns, the H pattern commonly occurred during early or late seizure propagation among 57 patients (81.4%). It harbored within specific EEG segments characterized by fast activity and irregular polyspikes. The H pattern often appeared simultaneously across different brain regions at a consistent fundamental frequency, highlighting a crucial stage in seizure propagation characterized by inter-regional synchronization. The *d*H pattern demonstrated greater nonlinearity compared to the non-*d*H pattern, as evidenced by bispectral analysis. The waveforms associated with the *d*H pattern were more stereotyped and showed increased skewness and/or asymmetry. Notably, the complete removal of areas exhibiting the *d*H pattern, but not high epileptogenicity index (≥0.3) or seizure onset zone, was independently associated with seizure freedom after surgery.

**Significance:** The H pattern provides unique insights into ictal neural dynamics. Additionally, it is a novel and alternative approach for measuring the EZ over an extended ictal time window.

**KEY POINTS:**

1. The harmonic pattern (H pattern) is commonly present in focal epileptic seizures and can help to improve the accuracy of EZ localization over an extended time window.
2. The H pattern is a spectral signature of waveform skewness or asymmetry. The dominant H pattern reflects a stronger nonlinearity of ictal EEG signals.
3. The H pattern can appear simultaneously in different areas with a consistent fundamental frequency, indicating a key stage of inter-regional synchronization.

## INTRODUCTION

Epilepsy is a chronic neurological disorder characterized by the tendency to have recurrent spontaneous seizures, which are associated with complex and dynamic patterns of neuronal synchrony and asynchrony. The accurate localization of the epileptogenic zone (EZ) is critical for achieving favorable surgical outcomes in drug-resistant focal epilepsy.^1,2^ Stereo-EEG (SEEG) is the most precise method for EZ identification. However, the ictal patterns observed on SEEG exhibit significant heterogeneity, suggesting a complex, highly dynamic, and individualized nature of the ictal network.^3,4^ The interpretation of these patterns primarily relies on visual inspection. The limited success rate of surgery, with only 50-70% of patients achieving long-term seizure freedom, is still considered unsatisfactory.^5^

To improve EZ delineation, numerous quantitative EEG analysis methods have been developed.^6^ Most approaches have focused on fast activity (FA, typically > 30 Hz), either as a standalone feature or in combination with other features.^7–13^ For example, epileptogenic index (EI) and epileptogenicity mapping measure the energy ratio changes between FA and slow wave activities during seizure onset, aiding in EZ localization.^7,8^ Emerging evidence highlights the importance of spectral architecture in ictal FA, not just its power.^14,15^ The ictal chirp, a high-power FA band (> 80 Hz) with a decremental frequency on time-frequency map over 5-10 seconds, is a common pattern that may assist in EZ localization.^14,16^ Grinenko et al. introduced EEG fingerprint as an ictal marker for EZ identification, noting that ictal FA consists of multiple narrow bands.^15,17^ Nevertheless, these methods demonstrate limited efficacy for slower seizure onset patterns, which account for 20-30% of cases,^18–20^ indicating the need for further investigation into ictal EEG spectral properties. Through comprehensive analysis of ictal SEEG spectral properties in focal epilepsy, we identified a unique spectral structure, referred to as the ‘harmonic pattern’ (H pattern), featuring multiple, equidistant, high-density frequency bands that vary over time (Figure 1). The H pattern is commonly observed in ictal EEG recordings from both experimental models of epilepsy and epileptic patients.^16,21,22^. Notably, the presence of harmonics in EEG or local field potential (LFP) suggests nonlinear neural oscillations, which are relevant to neural communication and brain dynamics.^23–25^ The H pattern may therefore provide straightforward insight into the nonlinear nature of the ictal networks. Here we systematically examined this phenomenon and evaluated its clinical significance in a cohort of patients with focal epilepsy undergoing SEEG evaluation. Furthermore, bispectral analysis was employed to interrogate the signal properties of the H pattern, thereby confirming its nonlinear nature.

**Figure 1.**
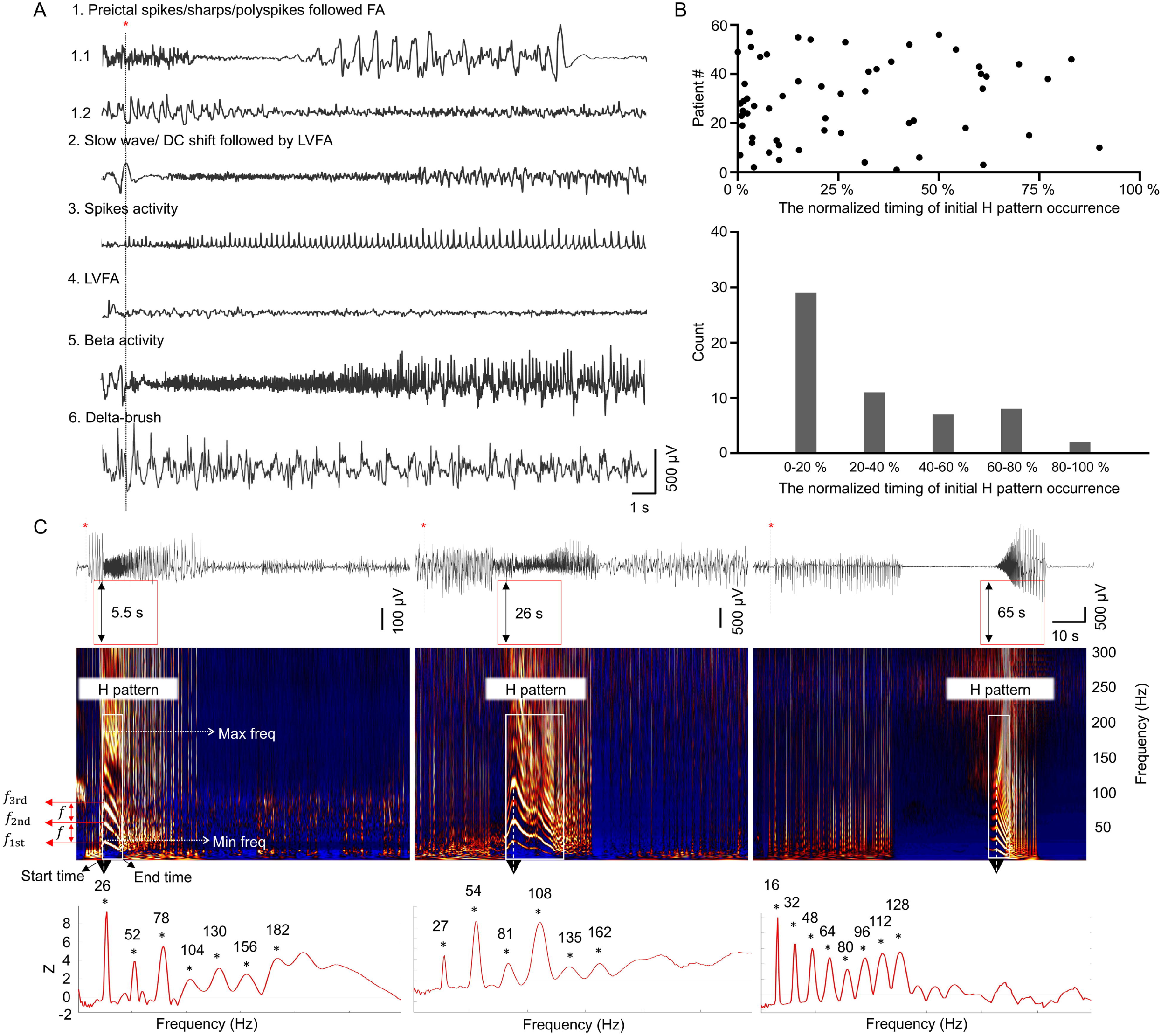
H patterns recorded in seizures with various ictal EEG onset patterns. **(A)** The six ictal onset patterns defined on SEEG. Pattern 1, preictal spikes/sharps/polyspikes following fast activity > 14 Hz (Pattern 1.1: LVFA, amplitude < 30 μV; Pattern 1.2: HVFA, amplitude >30 μV); Pattern 2, slow wave/DC shift followed by LVFA; Pattern 3, spikes activity; Pattern 4, LVFA; Pattern 5, beta activity; Pattern 6, delta-brush. **(B)** Distribution of the occurrence of the initial H pattern during seizures. Scatterplot showing the normalized timing of the initial H pattern start-point, expressed as a percentage of total seizure duration (top). Cumulative histogram showing the occurrence of the initial H pattern (bottom). **(C)** EEG (top) and corresponding TFM (middle) during seizures. The TFM shows the ‘harmonic pattern’ (H pattern): multiple equidistant high-density, narrow bands with varying frequency over time. The initial H patterns presented at various seizure stages on TFM. The black double arrows in the red box mark the start time point of the H pattern. The power spectral density corresponding to the maximal frequency of the H pattern (the black arrow) demonstrates an equidistant distribution of the frequency bands (bottom). The frequency intervals were 26, 27, and 16 Hz, respectively. LVFA: low-voltage fast activity; HVFA: high-voltage fast activity; TFM: time frequency map; Max freq: maximal frequency; Min freq: minimal frequency; *f*_1st_: the first frequency band, *f*_2nd_: the second frequency band, *f*_3rd_: the third frequency band; *f*: fundamental frequency (frequency interval). Red star: seizure onset.

## MATERIALS AND METHODS

### SEEG evaluation

This study was approved by the Medical Ethics Committee of our hospital. SEEG electrodes were implanted and reviewed in a bipolar montage (see supplementary methods). Only contacts localized within the grey matter were included in the analysis. The seizure onset pattern (SOP) was assessed based on the earliest electrophysiological changes recorded from the electrode contacts. The seizure onset zone (SOZ) was defined as the cortical region exhibiting the earliest ictal activity, while the early propagation zone (PZ) comprised areas showing ictal spread within 3-5 seconds of seizure onset. All other brain regions were classified as other zones (OZ).^12,26,27^ Two independent clinical neurophysiologists assessed SOP, SOZ, PZ, and OZ. Any discrepancies were resolved through discussion with a third epileptologist to reach a consensus.

Postoperative evaluation involved co-registration of preoperative MRI and postoperative CT scans to delineate resection boundaries. Electrode contacts within the resected area were classified as removed contacts. All patients were followed for a minimum of two years after surgery, with surgical outcomes assessed using the Engel’s classification, classifying as Engel Ia (seizure-free, SF) or > Engel Ia (not seizure-free, NSF).

### Quantitative EEG analysis

#### Identification of Harmonic patterns

For each seizure, a 110-second SEEG epoch (−10 s to +100 s relative to seizure onset) was extracted. Time-frequency maps (TFM) were generated using the Morlet wavelet transform, with a linear frequency resolution (1-300 Hz, 1 Hz step). Each TFM for each seizure was normalized using Z-score transformation, where power at each frequency was transformed by subtracting the mean and dividing by the standard deviation of the power over the entire 110-second window.

The H pattern was identified by its harmonic-like frequency distribution, consisting of multiple equidistant, high-density narrow bands that change over time (Figure 1C). These bands, from low to high frequencies, are labeled sequentially (e.g., *f*_1st_, *f*_2nd_, *f*_3rd_, etc.). The consistent difference between adjacent bands defines the frequency interval, which is equal to the fundamental frequency (*f*), typically the lowest band’s frequency. Narrow band frequencies align with multiples of *f*, with minimal and maximal frequencies marked by the first and last band peaks. Parameters of the H pattern, including minimal and maximal frequencies, and start and end times, were manually extracted using Brainstorm software. If multiple H patterns occurred sequentially within a single seizure, only the first one was analyzed.

To further identify contacts exhibiting dominant H pattern, a threshold was set at the upper quartile (Q3, 75th percentile) of the total number of bands:

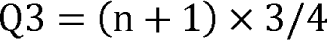

Where n is the highest band number in each seizure. Any contact with a band number of the H pattern exceeding this threshold would be designated as a ‘dominant-H pattern’ (*d*H pattern) contact; otherwise it was labeled as a ‘non-dominant H pattern’ (non-*d*H pattern) contact. Notably, the dominance of harmonic patterns is not determined by ictal discharge power, as substantial harmonics can occur with low discharge power or low EI (Figure S1).

#### Comparative analysis of the H pattern and other ictal EEG markers

We systematically assessed the diagnostic performance of the H pattern alongside established ictal EEG markers, including EI, SOZ, chirp, and EEG fingerprint. Chirp and EEG fingerprint were identified as previously described, and EI value was calculated using the AnyWave plugin.^7,14,15^ For EI analysis, we included only patients with ictal FA at onset and at least one contact with high EI (≥ 0.3). Since chirp and EEG fingerprint can be present across multiple brain regions and lack established thresholds for measuring epileptogenicity level, they were not included in quantitative analysis. For three markers (EI, SOZ and H pattern), the quantitative EZ (EZq) were defined as follows: contacts with high EI, contacts with SOZ, and contacts with *d*H pattern.

We evaluated the predictive performance of each marker using two complementary analyses. For EZ predictability assessment, using resected contacts from the SF patients as the reference standard, we classified contacts into four categories: true positives (TP, resected EZq), false positives (FP, non-resected EZq), true negatives (TN, non-resected non-EZq), and false negatives (FN, resected non-EZq). Sensitivity (TP/ (TP+FN)), specificity (TN/ (TN+FP)), precision (TP/ (TP+FP)), and Youden’s index (J = sensitivity + specificity - 1) were calculated for each marker. For outcome predictability analysis, patients were categorized based on the completeness of EZq resection (Yes/No) and surgical outcome (SF/NSF): TP (Yes, SF), FP (Yes, NSF), TN (No, NSF), and FN (No, SF). Receiver operating characteristic (ROC) curves were plotted, and the area under the curve (AUC) was calculated to assess the predictive performance for surgical outcomes. Additionally, we computed the resection ratio (number of resected EZq divided by total EZq),^26^ where a value of 1 indicates complete removal and 0 represents no removal.

#### Analysis of the signal properties of the H pattern

Harmonics can originate from either linear or nonlinear phenomena, each exhibiting their intrinsic characteristics (Figure 4A). Linear systems allow for the superposition of solutions, resulting in distinct harmonics. Conversely, nonlinear systems involve complex amplitude interactions that lead to phase correlations not detectable by spectral density alone. These phase correlations can be assessed through bispectral analysis (see supplementary methods; for details, refer to Sheremet et al., 2020).^24^ In brief, the bispectrum (including its normalized form) exhibits characteristic symmetries in the (*f_n_*, *f_m_*) plane (Figure 4B). The main diagonal serves as a symmetry axis.^28^ The red triangle highlights the nonredundant bispectral information, which is crucial for interpreting bispectral distributions. At any point (*f_n_*, *f_m_*), B (*f_n_*, *f_m_*) represents the phase correlation between the Fourier modes with *f_n_*, *f_m_* and *f_n+m_*. While the bispectrum is zero in linear systems with independent coefficients, nonlinear systems display peaks at phase-related triads, indicating two-wave coupling (Figure 4C).

To thoroughly analyze the nonlinear characteristics of the H pattern, we simultaneously measured the skewness and asymmetry of the waveforms. In the spectral domain, nonlinearity is expressed as the development of a chain of harmonics statistically phase coupled to fundamental frequency. In the time domain, it is expressed as the development of skewness and asymmetry in the waveform (Figure 4G-H). Skewness indicates horizontal asymmetry, where the duration of peaks or troughs is not half the period. Asymmetry indicates vertical asymmetry, with rise or decay times also not being half the period. Skewness and asymmetry can be extracted through bispectral analysis, as detailed in the supplementary methods.

#### Inferring seizure propagation

To investigate the propagation relationship between the *d*H pattern and the non-*d*H pattern, we utilized transfer functions. Specifically, for the propagation from the signal *a* to the signal *b*, the former was set as the input, and the latter as the fitting target, and *vice versa*. The fitting performance was evaluated by the optimal fitting percentage (FitPercent) among all possible transfer functions. FitPercent varies between -Infinity (bad fit) to 100 (perfect fit) (see supplementary methods for details). A higher FitPercent value indicates greater confidence in the propagation from signal *a* to signal *b*, implying that signal *a* likely contains sufficient information to reconstruct signal *b*.

### Statistical analysis

SPSS 24.0 was used for statistical analysis. Continuous variables were tested for homogeneity of variance and normality. Normal variables were presented as mean ± SD; non-normal variables were reported as medians with interquartile ranges. Categorical variables were displayed as frequencies. Two-group comparisons used Student’s t tests or Mann-Whitney U tests, and three-group analyses employed one-way ANOVAs or Kruskal-Wallis tests. Comparisons between two paired groups were performed using paired Student’s t-tests or Wilcoxon signed-rank tests. Pearson’s chi-square or Fisher’s exact tests were used for categorical variables. Multivariate logistic regression controlled for confounders in the *d*H pattern resection ratio comparisons. Significance was set at *P* < 0.05 (Bonferroni-adjusted for multiple testing).

## RESULTS

### Patients and SEEG evaluation

We retrospectively reviewed SEEG recordings from 80 patients with drug-resistant epilepsy (from January 2014 to April 2021). After excluding 3 patients with incomplete data and 7 with non-localizable SOZ, 70 patients were included in the final analysis. Of these, 57 (81.4%) patients presented with at least one H pattern, while the remaining 13 (18.6%) patients showed no discernible H pattern (Figure S2). The presence of H pattern was independent of SOP or presence of ictal fast activity (Figure 1A, Table S1), with no significant clinical differences between patients with versus without the H pattern. Notably,the H pattern was correlated with chirp and EEG fingerprint, but not with high EI (Table S1). Within those with H pattern, 39 (68.4%) achieved seizure freedom. No differences in various features were observed between the SF and NSF groups, except for number of electrodes and pathological findings (FCD II versus gliosis/nonspecific findings, Table S2).

### Classification of the H pattern

The initial H pattern emerged at 13.04 (22.41) s after seizure onset, occurring throughout seizure evolution (Figure 1B-C). The start time was not limited to the early stage of seizure evolution but occasionally extended to the late stage. Two distinct H pattern subtypes were identified: (1) the FA-H pattern (30/57, 52.6%), embedded in fast activity (> 25 Hz) and (2) the PS-H pattern (27/57, 47.4%), within irregular polyspikes (> 5 Hz). Compared to the PS-H pattern, the FA-H pattern displayed fewer frequency bands, a higher frequency interval, earlier start and end times, and greater minimal and maximal frequencies (Figure S3). Among all H patterns, the frequency bands dynamically evolved over time, displaying upward, downward, or bell-shaped trajectories before eventual disappearance. H patterns with downward or bell-shaped trajectories were highly concordant with chirp.

### Spatial distribution of the H pattern

The H pattern commonly occurred in the SOZ, PZ and sometimes in OZ at very close time points, displaying nearly identical frequency intervals, even when these regions were remotely separated. The H pattern’s end time and minimal/maximal frequencies did not vary across regions (Figure 2). Occasionally, the H pattern extended to the ipsilateral thalamus, appearing simultaneously with cortical regions and demonstrating the same frequency intervals (Figure S4). The proportion of contacts displaying the H pattern decreased from SOZ to PZ and then to OZ. Likewise, the number of bands exhibited a similar decreasing trend in each respective zone (Figure 2C). In subsequent analyses, to pinpoint H patterns that may indicate high epileptogenicity, we defined the *d*H pattern regions using Q3 for each patient.

**Figure 2.**
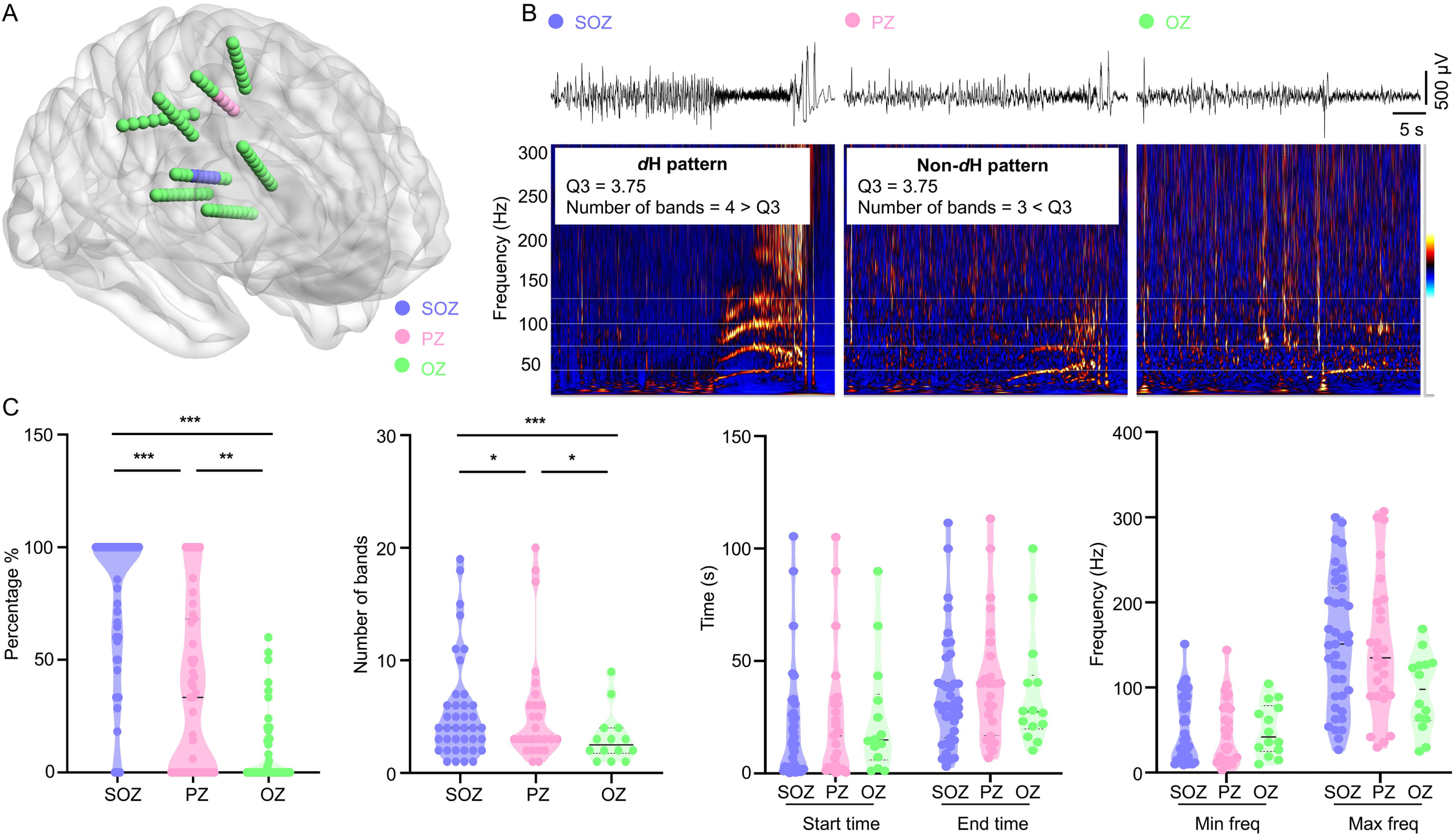
Distribution of the H pattern. **(A)** Anatomical distribution of SOZ, PZ, and OZ in a patient (left panel). In this case, SOZ is located in the precentral gyrus with early propagation to the postcentral gyrus. The right panel **(B)** shows the corresponding EEG recordings from SOZ, PZ, and OZ, along with the observed H pattern. **(C)** Comparison of parameters among SOZ, PZ, and OZ in group data. The distribution percentage and the number of bands of H pattern decrease in the order of SOZ, PZ, and OZ (Percentage %: SOZ: 100 (40), PZ: 33 (68), OZ: 0 (13), SOZ vs PZ, *P* < 0.001; SOZ vs OZ, *P* < 0.001; PZ vs OZ, *P* = 0.004. Number of bands: SOZ: 4 (4), PZ: 2 (4), OZ: 0 (2), SOZ vs PZ, *P* = 0.014; SOZ vs OZ, *P* < 0.001; PZ vs OZ, *P* = 0.012, nonparametric Kruskal-Wallis tests, after Bonferroni correction). \**P* < 0.05; \*\**P* < 0.01; \*\*\**P* < 0.001. Max freq: maximal frequency; Min freq: minimal frequency; SOZ: seizure onset zone; PZ: early propagation zone; OZ: other zone. *d*H pattern: dominant H pattern.

### Localization value of H pattern and other ictal markers

All 70 patients exhibited an identifiable SOZ. Most patients demonstrated the H pattern (81.4%), high EI (77.1%), and chirp (70%), while fewer exhibited the EEG fingerprint (35.7%) (Figure 3A). In terms of quantitative analysis, the overlap of EZq was used to assess concordance. Pairwise EZq overlap analysis revealed partial concordance between high EI and SOZ (31/54, 57.4%), *d*H pattern and SOZ (28/57, 49.1%), and high EI and *d*H pattern (21/45, 46.7%). Triple-marker analysis demonstrated partial overlap in 82.2% (37/45) of patients (Figure S5). No statistical difference was found in the number of EZq contacts among the three markers (Table S3).

**Figure 3.**
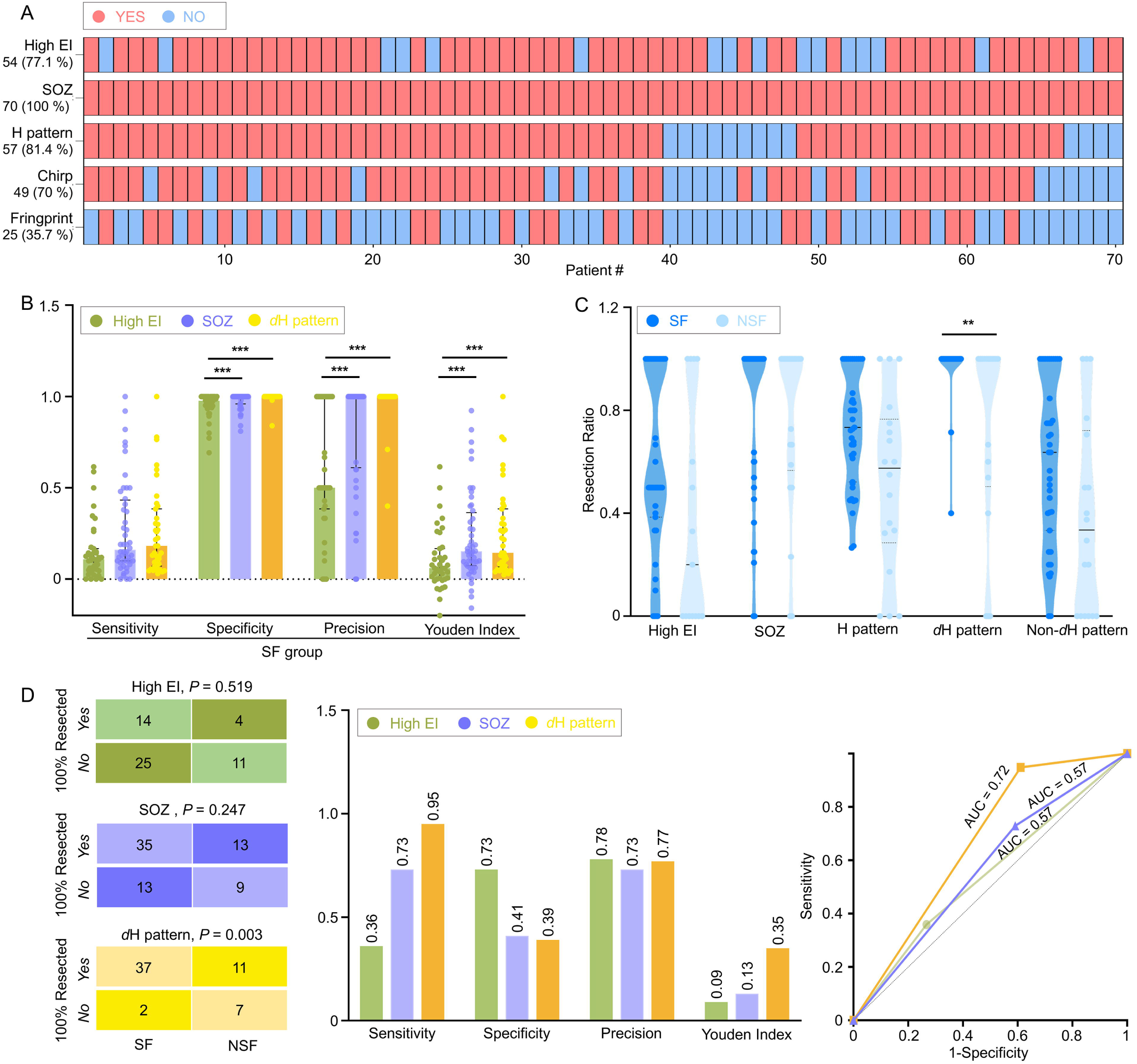
The localization value of the *d*H pattern. **(A)** The presence of five ictal markers in the 70 patients. Red = presence; blue = absence. For high EI, presence was defined by at least one contact with high EI; absence was defined by no high EI contacts or absence of FA at onset. **(B)** Statistics in predicting EZ in the SF group. Compared with high EI, the *d*H pattern and SOZ demonstrated higher specificity, precision and Youden index (Specificity: high EI: 0.98 (0.05), SOZ: 1.00 (0.04), *d*H pattern: 1 (0.00), high EI vs SOZ, *P* = 0.005; high EI vs *d*H pattern, *P* < 0.001; SOZ vs *d*H pattern, *P* = 0.057. Precision: high EI: 0.50 (0.62), SOZ: 1.00 (0.40), *d*H pattern: 1.0 (0.00), high EI vs SOZ, *P* = 0.001; high EI vs *d*H pattern, *P* < 0.001; SOZ vs *d*H pattern, *P* = 0.074. Youden index: high EI: 0.06 (0.17), SOZ: 0.15 (0.29), *d*H pattern: 0.14 (0.32), high EI vs SOZ, *P* = 0.006; high EI vs *d*H pattern, *P* < 0.001; SOZ vs *d*H pattern, *P* = 0.719. Nonparametric Kruskal-Wallis tests, after Bonferroni correction). **(C)** Resection ratio of the ictal markers in SF and NSF group. Only the resection ratio of the *d*H pattern differed significantly between the two groups (1.00 (0) vs. 1.00 (0.50), *P* = 0.005, nonparametric Mann-Whitney U test, after Bonferroni correction). **(D)** Statistics of ictal quantitative markers in predicting outcome. The *d*H pattern demonstrated superior predictive performance for seizure freedom, with the highest sensitivity (0.95), Youden index (0.35), and AUC (0.72; 95% CI: 0.57–0.84; *P* = 0.026) among three markers. \**P* < 0.05; \*\**P* < 0.01; \*\*\**P* < 0.001.

In predicting the EZ among SF patients, high EI demonstrated inferior specificity, precision, and Youden’s index compared to SOZ and *d*H pattern (Figure 3B). For outcome prediction, the *d*H pattern exhibited superior sensitivity of 0.95, and Youden’s index of 0.35. ROC analysis revealed that the *d*H pattern had the highest AUC (0.72; 95% CI: 0.57 - 0.84; *P* = 0.026), whereas the other two markers had lower and non-significant AUCs of 0.57 (both *P* > 0.05). However, no significant differences were found among the three AUCs according to the DeLong test (Figure 3D).

Furthermore, no difference in resection ratio was found except for *d*H pattern in the SF group compared to the NSF group (Figure 3C). On multivariate analysis, after adjusting for potential confounders of pathology, a complete resection of areas expressing the *d*H pattern was significantly associated with a higher chance of seizure freedom (OR = 32.33; 95% CI: 2.29 - 455.88; *P* = 0.01).

### Signal properties of the H pattern

The clinical implications of the H pattern were established in the above findings. To further investigate the factors contributing to its emergence, we analyzed the signal properties in both the frequency and time domains. In the frequency domain, bispectral analysis revealed that the emergence of the H pattern is predominantly due to nonlinear effects, with the *d*H pattern exhibited stronger nonlinear effects compared to the non-*d*H pattern (Figure 4D-F).

**Figure 4.**
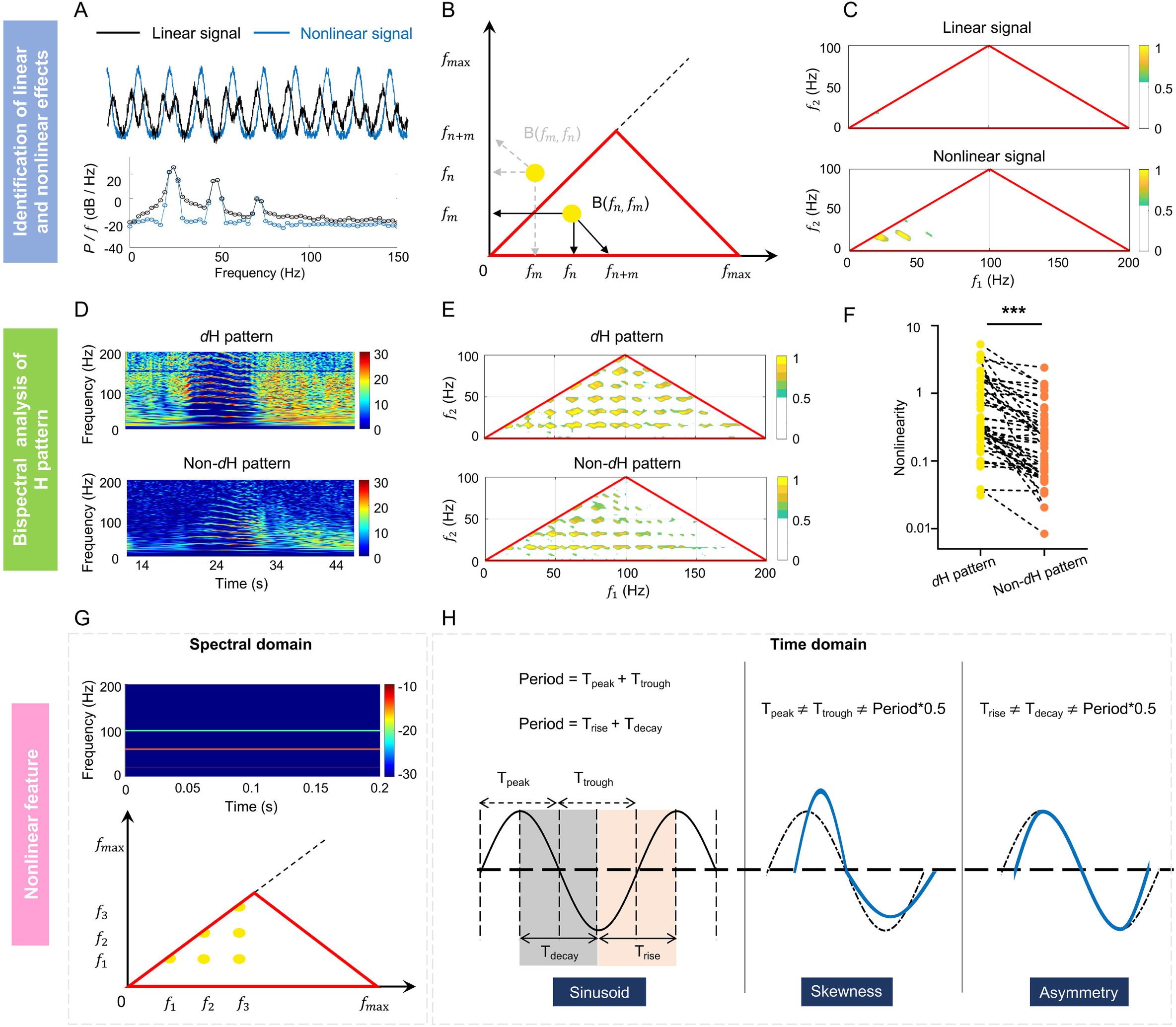
Analysis of the signal properties of the H pattern. ***Identification of linear and nonlinear effects:* (A)** Linear and nonlinear signals (top) and their PSD (bottom). PSD analysis cannot distinguish between linear and nonlinear time series, as these two oscillations have similar spectral density characteristics. **(B)** Bispectral symmetry. The main diagonal serves as the axis of symmetry, with identical data on both sides. The red triangle represents the area containing nonredundant bispectral information. The peak in the bispectral estimate (yellow dot) represents a phase-coupled triplet (*f_n_*, *f_m_*, *f_n+m_*). If a bispectral peak (yellow dot) is on the first diagonal, then m = n, meaning that frequency *f_n_* is phase correlated to its second harmonic *f*_2*n*_ = 2*f_n_*. **(C)** Normalized bispectrum of linear and nonlinear signals. Bispectral analysis can distinguish the linear and nonlinear characteristics of signals: the normalized bispectrum and all of its components for linear signal are approximately zero, whereas nonlinear signal exhibits four prominent peaks. ***Bispectral analysis of the H patterns:* (D)** Example of the *d*H and non-*d*H patterns. **(E)** Example of normalized bispectrum for the *d*H and non-*d*H patterns. In comparison to the non-*d*H pattern, the *d*H pattern exhibits more bicoherence peaks, signifying the presence of stronger harmonic phase coupling. **(F)** Comparison of bispectral values between the *d*H patterns and non-*d*H patterns in group data. The *d*H pattern showed stronger nonlinearity than the non-*d*H pattern (0.32 (0.9) vs 0.12 (0.28), *P* < 0.0001, nonparametric Mann-Whitney). ***Features of nonlinear effects:* (G)** In the spectral domain, nonlinearity is expressed as a series of harmonics that are phase-locked to the fundamental frequency, as evident in both TFM (top) and bispectral analysis (bottom). **(H)** In the time domain, nonlinearity is manifested as the asymmetry and skewness of the waveform. The left side uses a sinusoidal wave example to illustrate the period, peak, trough, rise, and decay. The distorted blue waveforms on the right, relative to the sinusoidal wave (black dashed line), exemplify skewness and asymmetry. PSD: power spectral density; DFT: discrete Fourier transform; TFM: time frequency map.

Nonlinearity was also evident in the time domain through waveform distortions, characterized by skewness and asymmetry (Figure 4G-H). In the following sections, we will carefully investigate the waveform characteristics of the EEG signals corresponding to the H pattern through four representative cases.

#### Stronger skewness underlying dH pattern

In the first case involving the FA-H pattern, the *d*H pattern displayed more distinct bands compared to the non-*d*H pattern, which was attributed to sharper troughs (Figure 5A-C). The *d*H pattern consistently exhibited shorter trough durations (T_trough_) than peak durations (T_peak_), contributing to a more stereotyped waveform morphology (Figure 5C). Bispectral analysis confirmed that the *d*H pattern had stronger bicoherence and skewness, indicative of sharper troughs, while these features were less pronounced in the non-*d*H pattern (Figure 5B). We constructed a simple waveform with sharper troughs, which exhibited pronounced harmonics. The corresponding bispectral analysis showed strong bicoherence and skewness, confirming that sharp troughs contribute to the emergence of the H pattern (Figure S6A-C, left).

**Figure 5.**
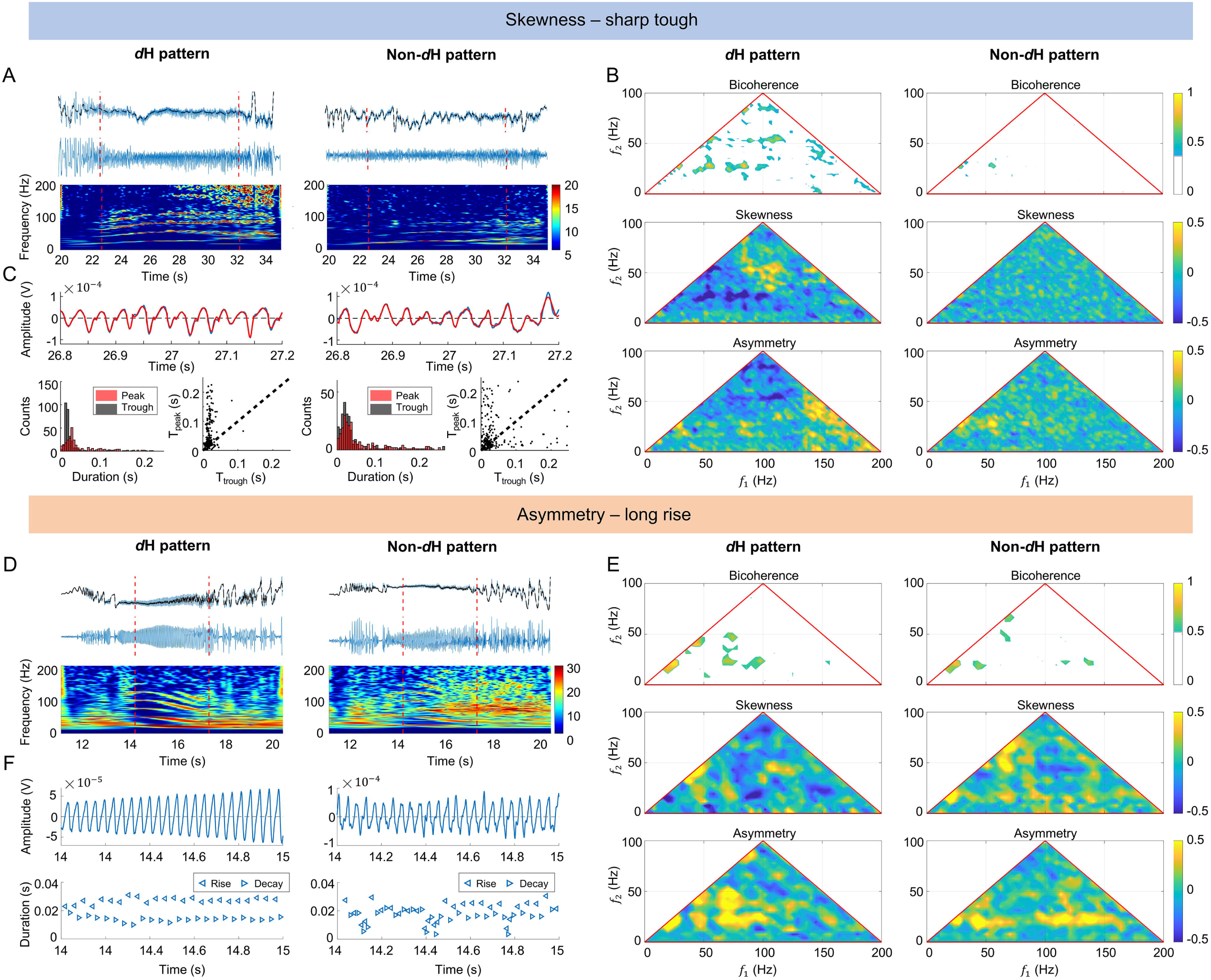
Waveform analysis of the H pattern. ***The dH pattern is attributed to stronger skewness (sharper trough).* (A)** Original SEEG signals in the *d*H/non-*d*H patterns (top), detrended SEEG signals (middle), and TFM (bottom); **(B)** Bispectral analysis for the *d*H/non-*d*H patterns. **(C)** Comparison of peak and trough. Top: peaks and troughs (blue lines) in the *d*H/non-*d*H patterns are separately fitted by sin waves (red lines), respectively; Bottom: Histogram (left) and scatter plot (right) depicting the distribution of T_peak_ vs. T_trough_ for the *d*H/non-*d*H patterns. ***The dH pattern is attributed to stronger asymmetry (longer rise).* (D)** Original SEEG signals in the *d*H/non-*d*H patterns (top), detrended SEEG signals (middle), TFM (bottom). **(E)** Bispectral analysis for the *d*H/non-*d*H patterns. **(F)** Comparison of rise and decay. Enlarged plots of detrended SEEG signals (top) and the corresponding rise (left triangles) and decay (right triangles) (bottom). The black dashed line in **A and D** represents the smoothed signals. The time with H pattern is marked by the red dotted vertical lines.

#### Stronger asymmetry underlying the dH pattern

In the second case with the FA-H pattern, the *d*H pattern was characterized by asymmetric waveforms, with rise durations (T_rise_) consistently longer than decay durations (T_decay_) (Figure 5D-F). This asymmetry contributed to a stereotyped EEG waveform. Bispectral analysis revealed stronger bicoherence and asymmetry in the *d*H pattern compared to the non-*d*H pattern (Figure 5E). A designed waveform with longer T_rise_ than T_decay_ also exhibited strong harmonics and bispectral characteristics, validating the role of asymmetric waveforms in the emergence of the H pattern (Figure S6A-C, right).

#### Stronger skewness and asymmetry underlying the dH pattern

The waveform associated with the H pattern sometimes concurrently manifested skewness and asymmetry. In one more case with an FA-H pattern, the *d*H pattern was characterized by sharp troughs and asymmetry (short rise, long decay) of waveforms (Figure S7A, C-D). The distorted waveforms consistently appeared for the *d*H pattern, causing a stereotyped morphology of the EEG segment. Bispectral analysis confirmed stronger bicoherence, skewness, and asymmetry in the *d*H pattern compared to the non-*d*H pattern (Figure S7B). Similar findings were observed in the PS-H pattern (supplementary results, Figure S8).

In summary, bispectral analysis revealed the existence of oscillatory interactions in the H pattern, suggesting a nonlinear effect. In the time domain, the H pattern was the spectral signature of the shape change of EEG waveforms.

### Propagation analysis of the H pattern

Focal seizures often originate from a localized region and subsequently propagate to other brain areas. Understanding this propagation is crucial for understanding seizure dynamics. Both *d*H and non-*d*H patterns typically share the same *f*, indicating synchronized oscillations between these regions and suggesting an underlying connection.

Nonlinear features of EEG signals can be influenced during the spatiotemporal propagation between regions. The stronger nonlinear effects observed in the *d*H pattern compared to the non-*d*H pattern may reflect the process of spatiotemporal propagation from the *d*H to the non-*d*H pattern. However, no significant differences were observed in the start and end times of the two patterns (Figure 6A). This may be attributed to the limited temporal resolution of the current measurements, which is insufficient to precisely resolve such rapid propagation processes. Therefore, we further evaluated this hypothesis by analyzing waveform similarity.

**Figure 6.**
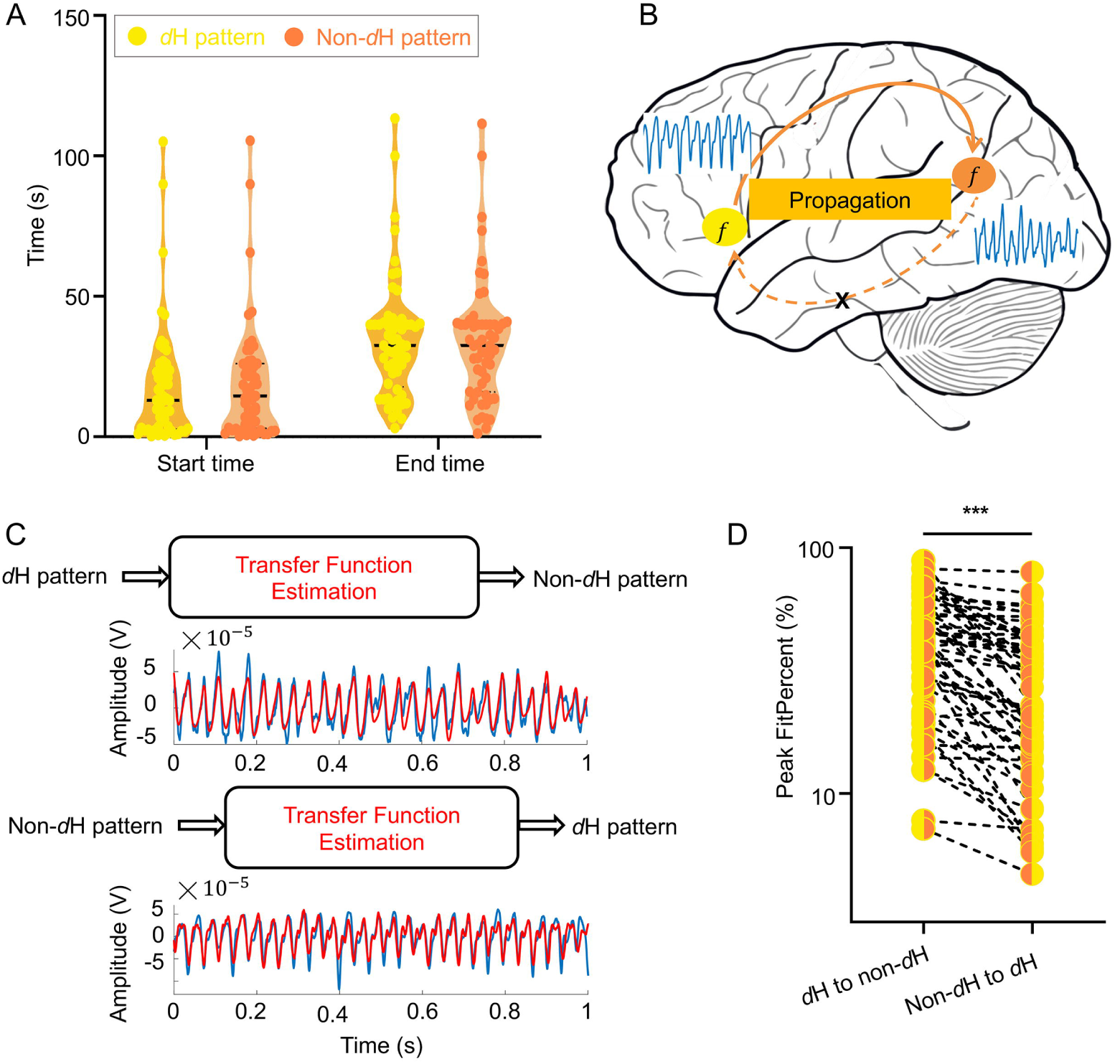
Propagation analysis of the H pattern. **(A)** Comparison of start and end times between the *d*H pattern and the non-*d*H pattern. **(B)** Schematic of signal propagation from region with the *d*H pattern to one with the non-*d*H pattern but not the opposite. The two regions share the same *f*. **(C)** Top: Estimating a transfer function with the input of *d*H pattern to fit non-*d*H pattern. Bottom: Estimating a transfer function with the input of non-*d*H pattern to fit *d*H pattern (blue: the input signal, red: the fitting signal). **(D)** Comparison of peak FitPercent between propagation from the *d*H pattern to the non-*d*H pattern and its opposite. The *d*H to non-*d*H direction showed a higher peak FitPercent (42.44 (34.44) vs 22.68 (28.72) %, *P* < 0.0001, Wilcoxon signed-rank). \**P* < 0.05; \*\**P* < 0.01; \*\*\**P* < 0.001. *f* : fundamental frequency.

We used transfer functions to evaluate the relationship between the *d*H and non-*d*H patterns. The results revealed that the peak FitPercent was higher when estimating propagation from the *d*H pattern to the non-*d*H pattern, compared to the reverse direction (Figure 6). This suggests that the *d*H pattern is more likely to propagate to the non-*d*H pattern, while propagation in the opposite direction occurs less frequently. This finding also implies that the *d*H pattern contains more information than the non-*d*H pattern.

## DISCUSSION

As a distinct spectral feature of ictal SEEG signals, the H pattern was commonly observed in most patients with various SOPs, whether early or late in seizure propagation. Its analysis not only provides crucial insights into the dynamics of ictal neural activity but also offers valuable information about the EZ. Our important findings include: (1) The dominance of H pattern is a promising and complementary ictal marker for identifying the EZ; (2) the H patterns represent spectral signature of waveform skewness and/or asymmetry, with the *d*H pattern reflecting a stronger nonlinearity of ictal oscillations, which correlates with higher epileptogenicity (Figure S9); (3) the H patterns can occur simultaneously in different cortical regions with consistent fundamental frequencies, suggesting they represent a pivotal stage of inter-regional synchronization during seizure propagation.

As a common ictal biomarker for focal epilepsy, the performance of ictal H pattern was non-inferior or better compared with that of high EI or SOZ in terms of EZ prediction and prognostic analysis. The resection ratio of the *d*H pattern was significantly higher in the SF group than NSF group. However, direct comparisons of AUC among the three quantitative markers revealed no significant differences, indicating that the predictive power of a single marker is limited. The presence of the H pattern did not change the overall surgical outcome; however, it may help EZ localization in the absence of other ictal markers. Integrating multiple ictal markers, together with interictal findings, is pertinent to improve EZ localization.

The FA-H and PS-H patterns manifested during different seizure stages. Compelling evidence indicates that the presence of fast activity is supported by a significant activation of inhibitory interneurons, combined with a transient shutdown of excitatory cells.^29–34^ Therefore, the FA-H pattern may be predominantly influenced by inhibitory interneurons. Irregular spiking usually occurs at the later stage of seizures and is linked to a decrease in inhibitory interneuron firing and an increase in excitatory neuron interactions.^35^ The PS-H pattern may be supported by a more substantial involvement of excitatory neuron firing.

Harmonic phenomena are prevalent in EEG/LFP signals, potentially linked to specific brain functional states.^36–38^ For instance, during non-rapid eye movement sleep, sleep spindles exhibit harmonic phenomena, with parameters changing in accordance with sleep stages, reflecting functional alterations in the thalamocortical reverberant network.^39^ Sheremet et al. studied the LFP recorded in the rat hippocampus and found harmonics of theta activity, together with the corresponding waveforms, changed with the movement speed. But harmonic phenomena have not received adequate attention in the field of epilepsy.^16,21,22^ Our findings reveal that harmonic features contain localizing information about the neural dynamics of seizures, enriching our understanding of the ictal network and underscoring the importance of considering its nonlinear aspects.

Neural oscillations are vital for understanding both normal and pathological brain states.^40,41^ However, traditional linear methods may miss the complexity of brain activity.^25^ Increasing evidence suggests that neural oscillations often have nonlinear properties.^24,25^ Our study shows that ictal signal nonlinearity, indicated by bispectral analysis and waveform skewness/asymmetry, is quantifiable, with the *d*H pattern exhibiting greater nonlinearity. Some studies showed nonlinear analysis of ictal EEG using sophisticated algorithms could provide seizure localization information.^12,42^ However, the nonlinear feature of the H pattern can be directly visualized in both the time and spectral domains, offering significant practical application value. In addition, our findings highlight the importance of waveform analysis, which can complement linear methods of EZ localization based on EEG frequency and power.

Many studies suggest that EEG/LFP waveforms can reflect the degree of correlated activity within neuronal populations. Sharp transients observed in spike-wave discharges are indicative of synchronous neuronal firing.^43^ Recent research has shown that increased neuronal synchronization results in highly asymmetric waveforms, while reduced synchronization leads to more sinusoidal shapes.^44^ Additionally, a stronger nonlinear component in the waveform often indicates greater neuronal synchronization in the cerebral cortex.^45^ Our results show that the *d*H pattern exhibits greater skewness and/or asymmetry in its waveforms, with a more stereotyped waveform. This consistency suggests the repeated occurrence of the same oscillatory pattern. These findings imply that the H pattern may serve as an EEG signature of rhythmic and synchronized neuronal firing within local epileptic tissues (Figure S9). Further research is imperative to elucidate the cellular and dynamic processes responsible for the diverse waveforms associated with the H pattern.

The H pattern represents a distinctive stage in seizure propagation. Our findings indicate a higher likelihood that signals in regions exhibiting the *d*H pattern propagate to the non-*d*H pattern, aligning with the gradient of epileptogenicity. The H pattern visually represents this unique stage of inter-regional synchronization, as evidenced by the consistent *f* observed across regions. This synchronization could be a characteristic of seizure propagation and can reach distant areas, such as the contralateral cortex and ipsilateral thalamus.

The H pattern can be observed throughout the entire seizure phase and is closely associated with the rhythmic firing and dynamic synchronization of neurons in epileptic tissues. This finding is consistent with prior research indicating that neuronal synchronization during seizures is a dynamic process, characterized by alternating phases of synchronization and desynchronization, reaching its peak at seizure termination.^46–48^ The presence of multiple H patterns during some seizures likely reflects the dynamic fluctuations in synchronization, suggesting a dynamic change in synchronization throughout the seizure which required further investigation.

The study has several limitations. First, due to the heterogeneous morphology of H patterns across subjects and the current lack of automated identification, their visual recognition was inherently subjective to personal experience. Second, simultaneous unit recording was not performed, which limits the exploration of the cellular mechanisms underlying the H pattern. Moreover, multiple H patterns sometimes occur in the same seizure, but only the first occurrence was analyzed in this study. Further investigation is warranted for cases where H patterns appear consecutively. Third, using a threshold (Q3) to define the dominant H pattern may not accurately capture individual variability. Future studies could benefit from adopting a patient-specific approach to defining the dominant H pattern. Lastly, we did not identify clinical factors association with presence of H pattern, which warrant further investigation in a larger cohort. Additionally, spatial sampling remains an intrinsic limitation of any SEEG study.

Our study defines a common and distinctive ictal spectral feature, termed the H pattern. Its dominance signifies specific waveforms and highlights the nonlinearity of ictal EEG signals. This feature may be associated with rhythmic and synchronized neuronal firing in local epileptic tissues. The H pattern imparts unique information about ictal neural dynamics as well as offers novel insights into EZ localization. Our data also provides evidence supporting an elongated time window for measuring EZ using quantitative EEG.

## Acknowledgement statement

This work was supported by the National Natural Science Foundation of China (grant numbers: 82171437, 82471469 and 82301636) and the Natural Science Foundation of Zhejiang Province (grant numbers: LD24H090003 and LY24H090004).

## Author contributions

Conceptualization: S.W., D.-P.Y. Data collection: L.-L.H., L.-Q.Y., H.-Y.Y. Data analysis: L.-L.H., X.-C.L., K.X., Y.-M.Z. Validation of analysis: Z.Z., H.-J.J. Supervision: Z.-J.W., Y.-C.W., K.-J.H. Funding acquisition: S.W., C.C. Writing-manuscript preparation: L.-L.H., D.-P.Y., J.M.Z. Writing-reviewing and editing: S.W., D.-P.Y., J.-P.Z., M.-P.D., C.Z.

## Data availability

The collected EEG data is used for seizure localization and to inform clinical decisions. The de-identified dataset supporting the study findings is available upon request from the corresponding author. For inquiries or to request the source data for the figures, please contact S.W. via email. EEG data analyses were performed using the freely available toolbox Brainstorm in combination with custom Matlab scripts, which are available at: https://github.com/chenx-epi/H-pattern.

## Supplementary Materials

Supplementary material is available online.

---

**Table S1.**
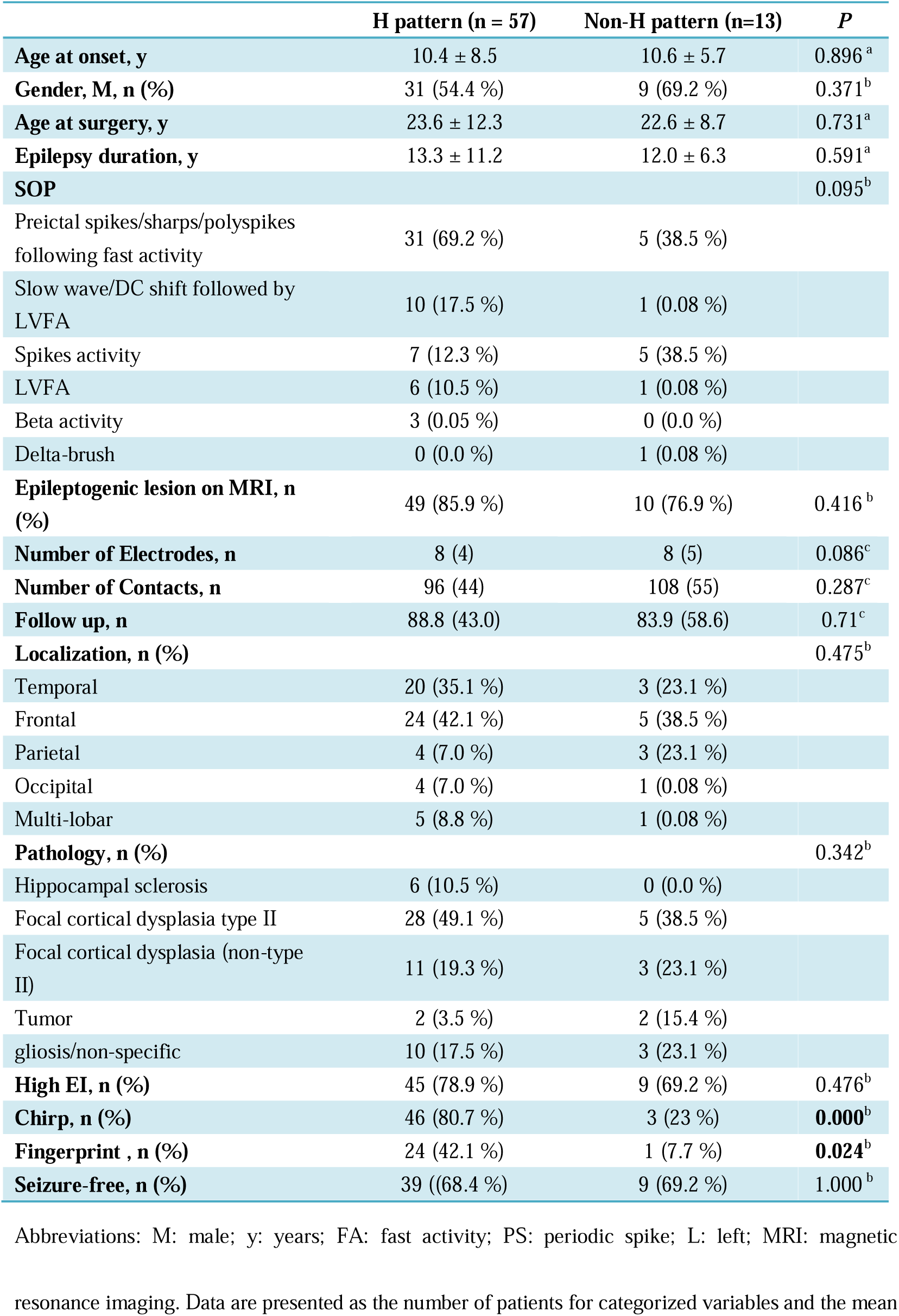
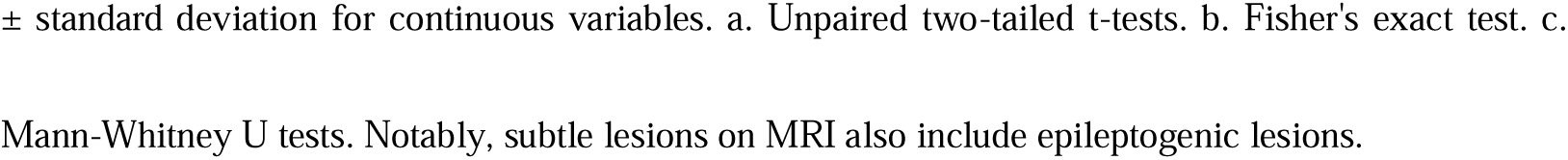
Clinical characteristics and surgical outcomes of patients with and without H pattern.

|  | H pattern (n = 57) | Non-H pattern (n=13) | P |
| --- | --- | --- | --- |
| **Age at onset, y** | 10.4 ± 8.5 | 10.6 ± 5.7 | 0.896<sup>a</sup> |
| **Gender, M, n (%)** | 31 (54.4 %) | 9 (69.2 %) | 0.371<sup>b</sup> |
| **Age at surgery, y** | 23.6 ± 12.3 | 22.6 ± 8.7 | 0.731<sup>a</sup> |
| **Epilepsy duration, y** | 13.3 ± 11.2 | 12.0 ± 6.3 | 0.591<sup>a</sup> |
| **SOP** |  |  | 0.095<sup>b</sup> |
| Preictal spikes/sharps/polyspikes following fast activity | 31 (69.2 %) | 5 (38.5 %) |  |
| Slow wave/DC shift followed by LVFA | 10 (17.5 %) | 1 (0.08 %) |  |
| Spikes activity | 7 (12.3 %) | 5 (38.5 %) |  |
| LVFA | 6 (10.5 %) | 1 (0.08 %) |  |
| Beta activity | 3 (0.05 %) | 0 (0.0 %) |  |
| Delta-brush | 0 (0.0 %) | 1 (0.08 %) |  |
| **Epileptogenic lesion on MRI, n (%)** | 49 (85.9 %) | 10 (76.9 %) | 0.416<sup>b</sup> |
| **Number of Electrodes, n** | 8 (4) | 8 (5) | 0.086<sup>c</sup> |
| **Number of Contacts, n** | 96 (44) | 108 (55) | 0.287<sup>c</sup> |
| **Follow up, n** | 88.8 (43.0) | 83.9 (58.6) | 0.71<sup>c</sup> |
| **Localization, n (%)** |  |  | 0.475<sup>b</sup> |
| Temporal | 20 (35.1 %) | 3 (23.1 %) |  |
| Frontal | 24 (42.1 %) | 5 (38.5 %) |  |
| Parietal | 4 (7.0 %) | 3 (23.1 %) |  |
| Occipital | 4 (7.0 %) | 1 (0.08 %) |  |
| Multi-lobar | 5 (8.8 %) | 1 (0.08 %) |  |
| **Pathology, n (%)** |  |  | 0.342<sup>b</sup> |
| Hippocampal sclerosis | 6 (10.5 %) | 0 (0.0 %) |  |
| Focal cortical dysplasia type II | 28 (49.1 %) | 5 (38.5 %) |  |
| Focal cortical dysplasia (non-type II) | 11 (19.3 %) | 3 (23.1 %) |  |
| Tumor | 2 (3.5 %) | 2 (15.4 %) |  |
| gliosis/non-specific | 10 (17.5 %) | 3 (23.1 %) |  |
| **High EI, n (%)** | 45 (78.9 %) | 9 (69.2 %) | 0.476<sup>b</sup> |
| **Chirp, n (%)** | 46 (80.7 %) | 3 (23 %) | **0.000<sup>b</sup>** |
| **Fingerprint, n (%)** | 24 (42.1 %) | 1 (7.7 %) | **0.024<sup>b</sup>** |
| **Seizure-free, n (%)** | 39 ((68.4 %) | 9 (69.2 %) | 1.000<sup>b</sup> |

Abbreviations: M: male; y: years; FA: fast activity; PS: periodic spike; L: left; MRI: magnetic resonance imaging. Data are presented as the number of patients for categorized variables and the mean
± standard deviation for continuous variables. a. Unpaired two-tailed t-tests. b. Fisher's exact test. c.
Mann-Whitney U tests. Notably, subtle lesions on MRI also include epileptogenic lesions.

**Table S2.**
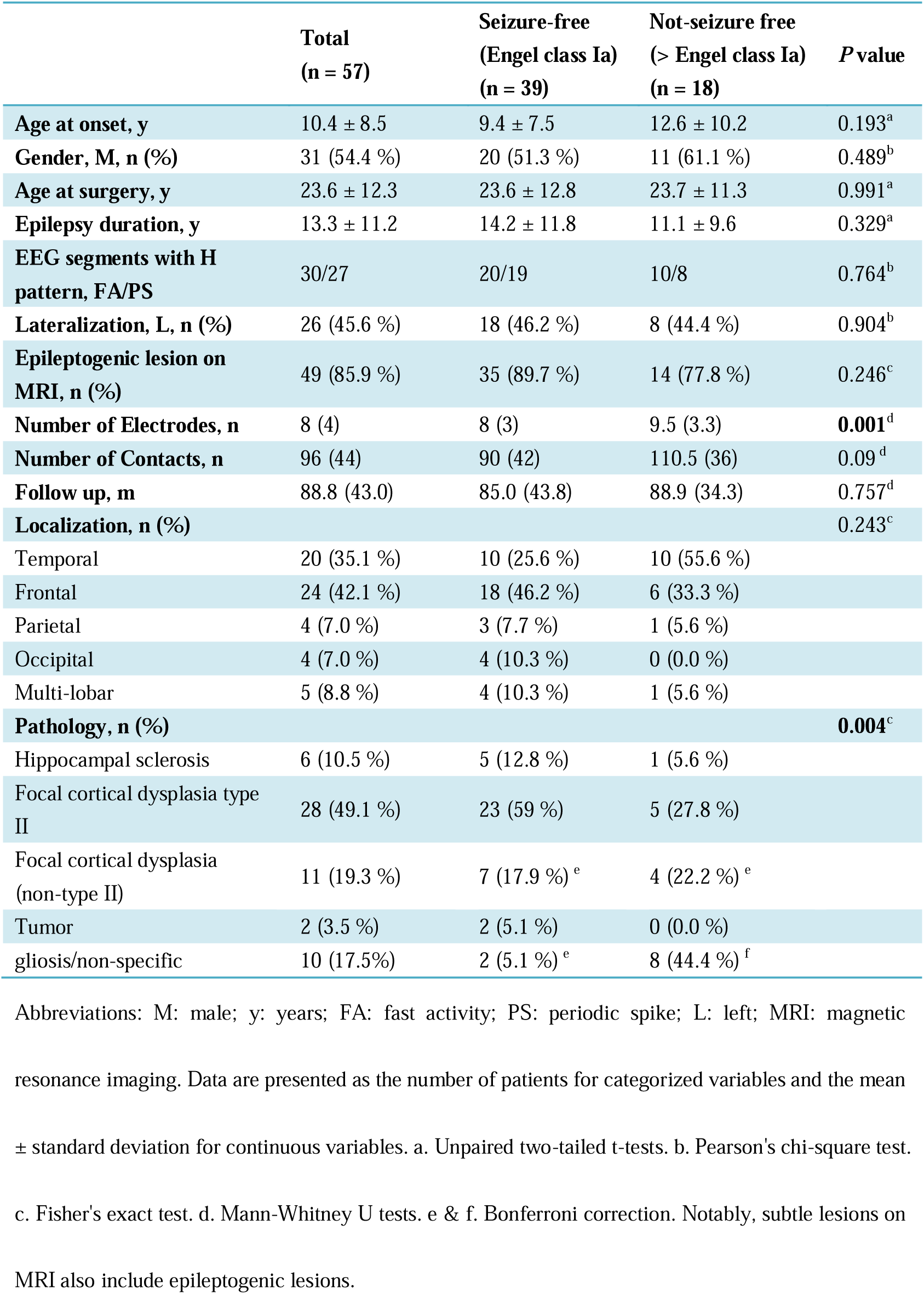
Clinical characteristics comparison of patients with H pattern by prognosis.

|  | **Total**<br>**(n = 57)** | **Seizure-free**<br>**(Engel class Ia)**<br>**(n = 39)** | **Not-seizure free**<br>**(> Engel class Ia)**<br>**(n = 18)** | **P value** |
| --- | --- | --- | --- | --- |
| **Age at onset, y** | 10.4 ± 8.5 | 9.4 ± 7.5 | 12.6 ± 10.2 | 0.193<sup>a</sup> |
| **Gender, M, n (%)** | 31 (54.4 %) | 20 (51.3 %) | 11 (61.1 %) | 0.489<sup>b</sup> |
| **Age at surgery, y** | 23.6 ± 12.3 | 23.6 ± 12.8 | 23.7 ± 11.3 | 0.991<sup>a</sup> |
| **Epilepsy duration, y** | 13.3 ± 11.2 | 14.2 ± 11.8 | 11.1 ± 9.6 | 0.329<sup>a</sup> |
| **EEG segments with H**<br>**pattern, FA/PS** | 30/27 | 20/19 | 10/8 | 0.764<sup>b</sup> |
| **Lateralization, L, n (%)** | 26 (45.6 %) | 18 (46.2 %) | 8 (44.4 %) | 0.904<sup>b</sup> |
| **Epileptogenic lesion on**<br>**MRI, n (%)** | 49 (85.9 %) | 35 (89.7 %) | 14 (77.8 %) | 0.246<sup>c</sup> |
| **Number of Electrodes, n** | 8 (4) | 8 (3) | 9.5 (3.3) | **0.001<sup>d</sup>** |
| **Number of Contacts, n** | 96 (44) | 90 (42) | 110.5 (36) | 0.09<sup>d</sup> |
| **Follow up, m** | 88.8 (43.0) | 85.0 (43.8) | 88.9 (34.3) | 0.757<sup>d</sup> |
| **Localization, n (%)** |  |  |  | 0.243<sup>c</sup> |
| Temporal | 20 (35.1 %) | 10 (25.6 %) | 10 (55.6 %) |  |
| Frontal | 24 (42.1 %) | 18 (46.2 %) | 6 (33.3 %) |  |
| Parietal | 4 (7.0 %) | 3 (7.7 %) | 1 (5.6 %) |  |
| Occipital | 4 (7.0 %) | 4 (10.3 %) | 0 (0.0 %) |  |
| Multi-lobar | 5 (8.8 %) | 4 (10.3 %) | 1 (5.6 %) |  |
| **Pathology, n (%)** |  |  |  | **0.004<sup>c</sup>** |
| Hippocampal sclerosis | 6 (10.5 %) | 5 (12.8 %) | 1 (5.6 %) |  |
| Focal cortical dysplasia type<br>II | 28 (49.1 %) | 23 (59 %) | 5 (27.8 %) |  |
| Focal cortical dysplasia<br>(non-type II) | 11 (19.3 %) | 7 (17.9 %)<sup>e</sup> | 4 (22.2 %)<sup>e</sup> |  |
| Tumor | 2 (3.5 %) | 2 (5.1 %) | 0 (0.0 %) |  |
| gliosis/non-specific | 10 (17.5%) | 2 (5.1 %)<sup>e</sup> | 8 (44.4 %)<sup>f</sup> |  |

Abbreviations: M: male; y: years; FA: fast activity; PS: periodic spike; L: left; MRI: magnetic resonance imaging. Data are presented as the number of patients for categorized variables and the mean ± standard deviation for continuous variables. a. Unpaired two-tailed t-tests. b. Pearson's chi-square test. c. Fisher's exact test. d. Mann-Whitney U tests. e & f. Bonferroni correction. Notably, subtle lesions on MRI also include epileptogenic lesions.

**Table S3.** Comparison of contact numbers for the three markers.

| Number of Contacts, n | High EI | SOZ | dH pattern | *P* |
| --- | --- | --- | --- | --- |
| **Total** | 4 (3.25) | 5 (7.5) | 4 (5.5) | 0.062 |
| **Seizure-free group ( SF )** | 4 (3) | 5 (8) | 5 (6) | 0.379 |
| **Not-seizure free group ( NSF )** | 4 (4) | 6 (7.5) | 3 (5) | 0.105 |
| ***P* ( SF vs NSF )** | 0.382 | 0.604 | 0.829 |  |

