## Supplementary Materials for "Harmonic patterns embedded in ictal EEG signals in focal epilepsy: new insight into the epileptogenic zone"

Lingli Hu, *et al.*

Dongping Yang..

**Supplementary Text**

**Supplementary Materials and Methods**

**SEEG evaluation**

We conducted a review of consecutive patients with drug-resistant epilepsy who underwent SEEG evaluation followed by resective surgery at our epilepsy center. A comprehensive assessment was undertaken, encompassing a detailed medical history, neuropsychological testing, neuroimaging examination, and scalp video-EEG recordings. SEEG was performed using intracerebral multiple-contact electrodes, with contacts that were 2 mm in length, 0.8 mm in diameter, and 1.5 mm apart. SEEG signal was recorded by EEG systems with 256 channels (Nihon Kohden) or 128 channels (Xltek, Natus, Ontario, Canada) at a sampling rate of 1000 or 2000 Hz. Pre-implantation MRI and post-implantation CT were co-registered to locate each contact anatomically along each electrode trajectory.

**Bispectral analysis**

The basic formula of bispectral analysis is as follows:

$$B_{n,m}=B\left( f_{n},f_{m} \right)=E[G_{n}G_{m}G_{m+n}^{*}]$$

Here,$f_{n}$ and $f_{m}$ are two frequencies, $G_{n}$ is the corresponding complex Fourier coefficient using the discrete Fourier transform (DFT), and * denotes the complex conjugate. The bispectrum $B_{n,m}$ represents the phase correlation between Fourier modes with frequencies $f_{n}, f_{m}$ and $f_{n+m}=f_{n}+f_{m}$.

To eliminate the distortion induced by the variance distribution, the bispectrum can be normalized as follows:

$b_{n,m}=\frac{B_{n,m}}{\left( E[\left| G_{n}G_{m} \right|^{2}]E[\left| G_{n+m} \right|^{2}] \right)^{1/2}}$.

The squared modulus and phase of the normalized bispectrum $b_{n,m}$ are termed bicoherence and biphase, respectively.^1^

The normalized bispectrum's real and imaginary parts encapsulate the "frequency distribution" of these waveform distortions. We will refer to the real and imaginary parts of the normalized bispectrum as the distributions of skewness and asymmetry, defined as follows:

$\xi_{n,m}=Re\{b_{n,m}\}$; $A_{n,m}=Im\{b_{n,m}\}$

Here, Re{z} and Im{z} denote the real and imaginary parts of the complex number z, respectively. The sign of $\xi$ and $A$ ($\pm$) indicates the direction of the skewness or asymmetry.

Furthermore, waveform original SEEG signals were carefully analyzed. Initially, signals were detrended using the smoothing method 'loess' in MATLAB, which adjusted the wave's oscillations around a zero-valued baseline, thereby distinguishing troughs from peaks. Each trough and peak was subsequently fitted with a sinusoidal wave of a single frequency to identify the waveforms of EEG signals.

EEG signals were processed by de-meaning, linear detrending, and dividing into 50% overlapping segments of approximately 0.5-second windows ($2^{10}$-point windows at a sampling rate $f_{s}=2000$ Hz), achieving a frequency resolution of approximately $\Delta f=2$ Hz. The total intervals were around 7 seconds, yielding approximately 28 degrees of freedom (DOF). For the normalization $b_{n,m}$, the probability density function was approximated using the noncentral $\chi^{2}$ distribution.^2,3^ A confidence level distinguishes linear from nonlinear stochastic processes. With DOF = 28, zero-mean bicoherence, significant in this context, is $\left| b \right|<\sqrt{\frac{6}{DOF}}\approx0.46$ for a 95% confidence level.^2,4^ All calculations were implemented in MATLAB, utilizing its DFT functions. Bispectral estimates were computed using code based on modified functions from the HOSA toolbox.

**Inferring seizure propagation**

The formula for FitPercent is as follows:

$$\mathrm{FitPercent}=100\left( 1-\frac{\left\| y_{\mathrm{measured}}-y_{\mathrm{model}} \right\|}{\left\| y_{\mathrm{measured}}-\bar{y_{\mathrm{measured}}} \right\|} \right)$$

Where $y_{\mathrm{measured}}$ is the target output data, $\bar{y_{\mathrm{measured}}}$ is its mean, and $y_{\mathrm{model}}$ is the simulated or predicted response of the model. $\left\| * \right\|$ indicates the 2-norm of a vector.

**Supplementary Results**

**Stronger skewness and asymmetry underlying the *d*H pattern**

We presented a case with the PS-H pattern (Fig. S8). This pattern was characterized by both sharp peaks and asymmetric (short rise, long decay) waveforms (Fig. S8A, C, D). The highly stereotyped waveform of the *d*H pattern was attributed to its consistently sharp peaks and asymmetric pattern. Similarly, we found that the *d*H pattern exhibited stronger bicoherence, skewness, and asymmetry compared to the non-*d*H pattern (Fig. S8B).

**Supplementary Figures**

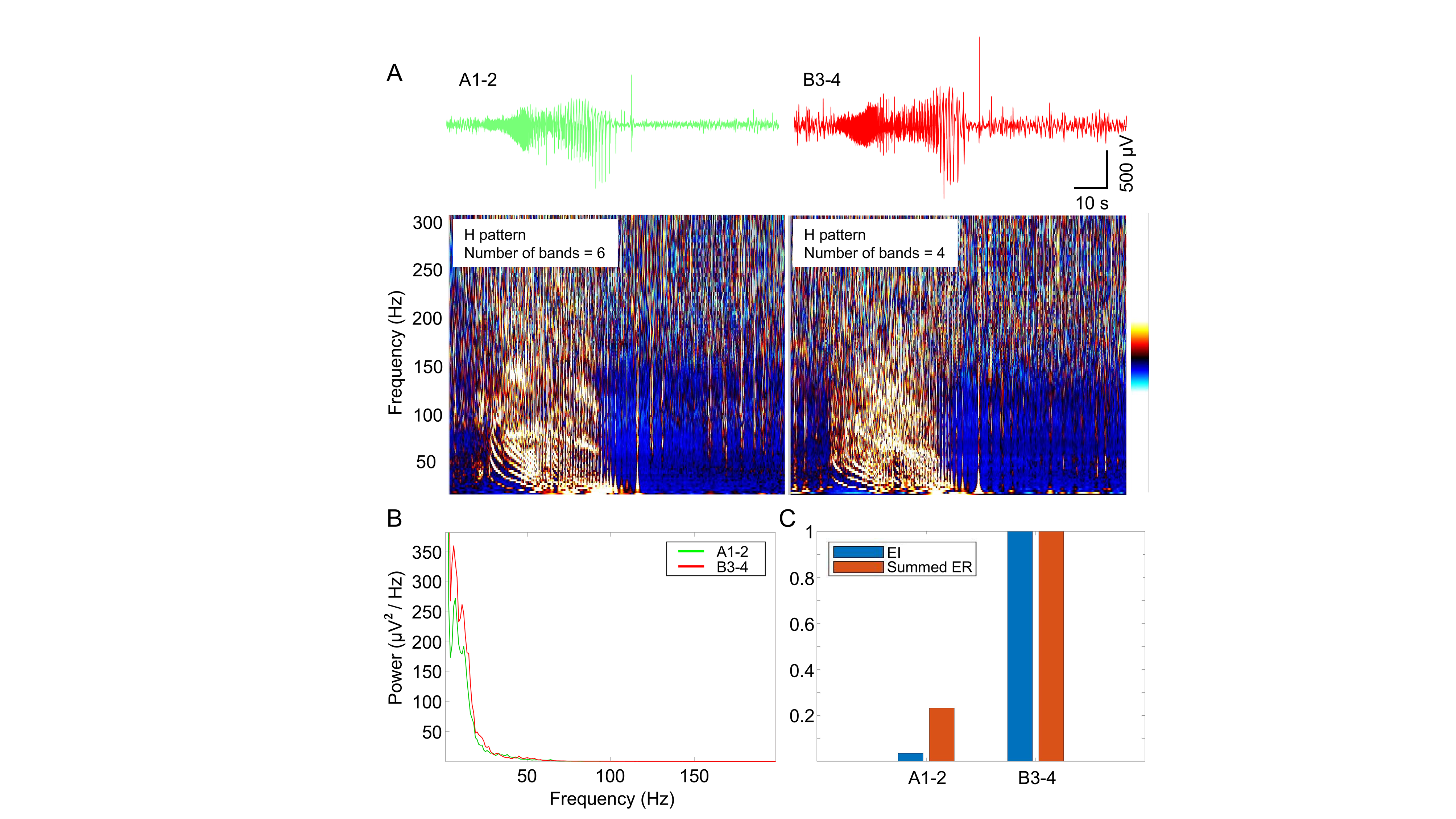

**Supplementary Figure 1 Representative illustration of the relationship between ictal discharge power and harmonic patterns.** **(A)** EEG recording (top) and corresponding time-frequency map (bottom) during a seizure. **(B)** Corresponding power spectral density. **(C)** Corresponding EI analysis. A1-2 exhibited a higher number of H pattern bands than B3-4, despite having lower ictal discharge power and EI values.

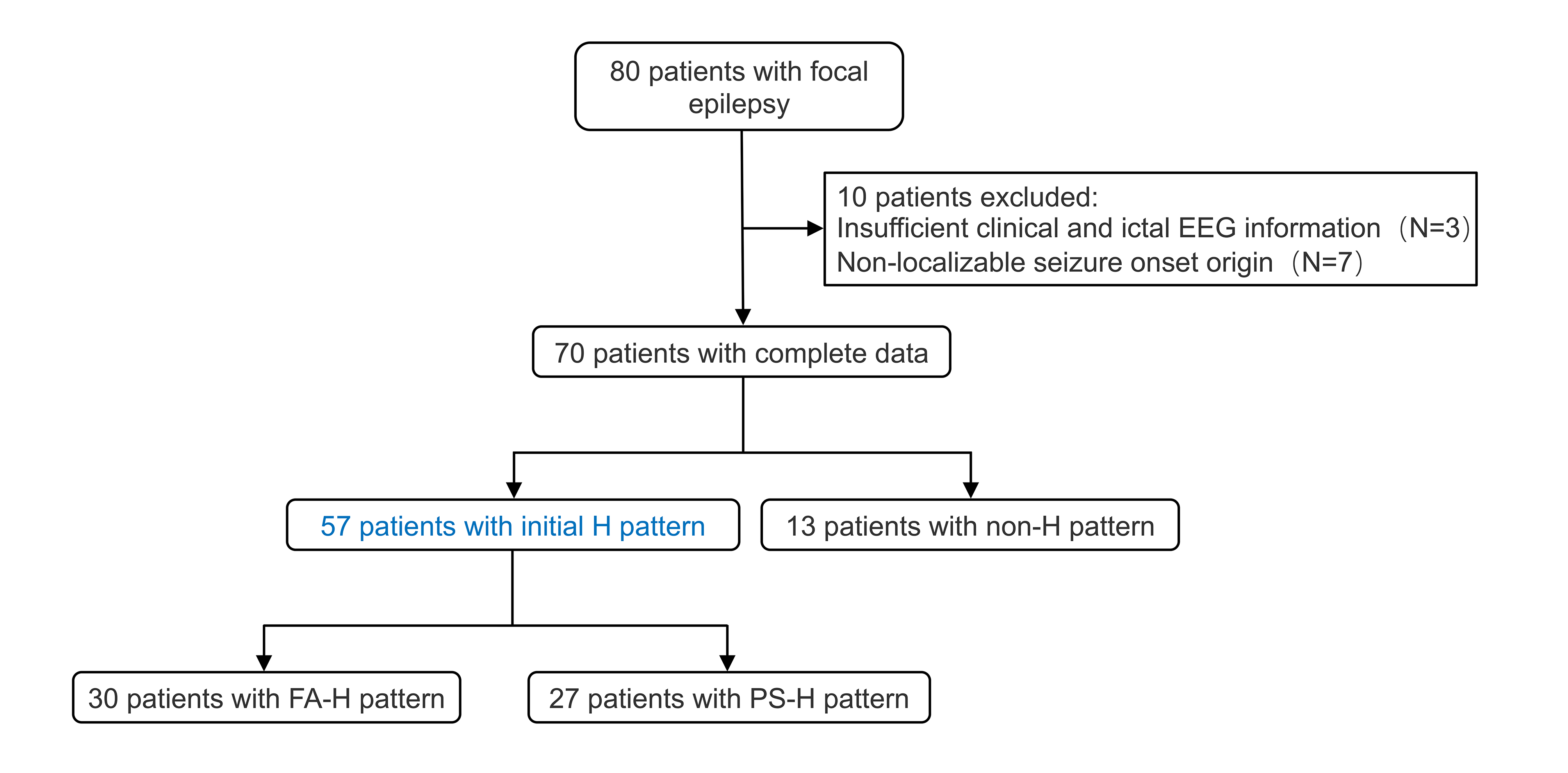

**Supplementary Figure 2 Flow chart showing the inclusion and exclusion process of patients in our study.**

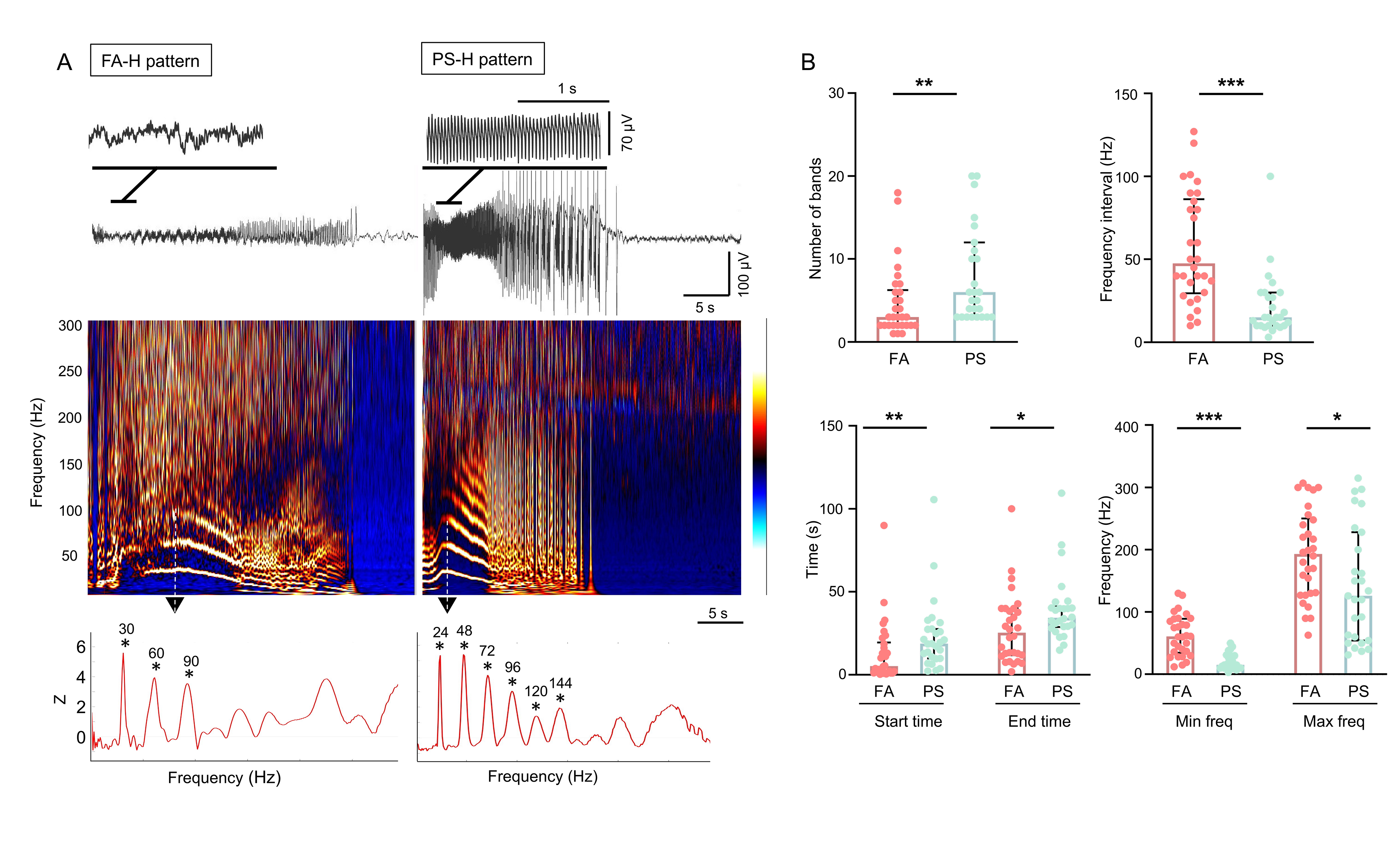

**Supplementary Figure 3 Two types of EEG segments harboring H pattern. (A)** H pattern is presented in fast activity (FA-H pattern) and irregular polyspikes (PS-H pattern) (top and middle). The power spectral density (bottom) at the maximal frequency point shows an equidistant distribution of the frequency bands. **(B)** Comparison of parameters between the FA-H pattern and PS-H pattern. The former showed a fewer number of frequency bands (3 (4) vs. 6 (8), *P* = 0.006), higher frequency interval (47.5 (56.75) vs. 15 (20.00) Hz, *P* < 0.0001), earlier start (time zero, EEG onset time; 5.2 (18.19) vs. 18.70 (17.20) s, *P* = 0.004) and end (25.4 (28.30) vs. 33.9 (11.70) s, *P* = 0.015) time, and higher minimal (60 (54.70) vs. 15.00(20.00) Hz, *P* < 0.0001) and maximal (193.00 (120.75) vs. 126.00 (159.00) Hz, *P* = 0.021) frequencies than the latter. **P* < 0.05; ***P* < 0.01; ****P* < 0.001. Max freq: maximal frequency; Min freq: minimal frequency. Statistical analysis was performed using the nonparametric Mann-Whitney U test.

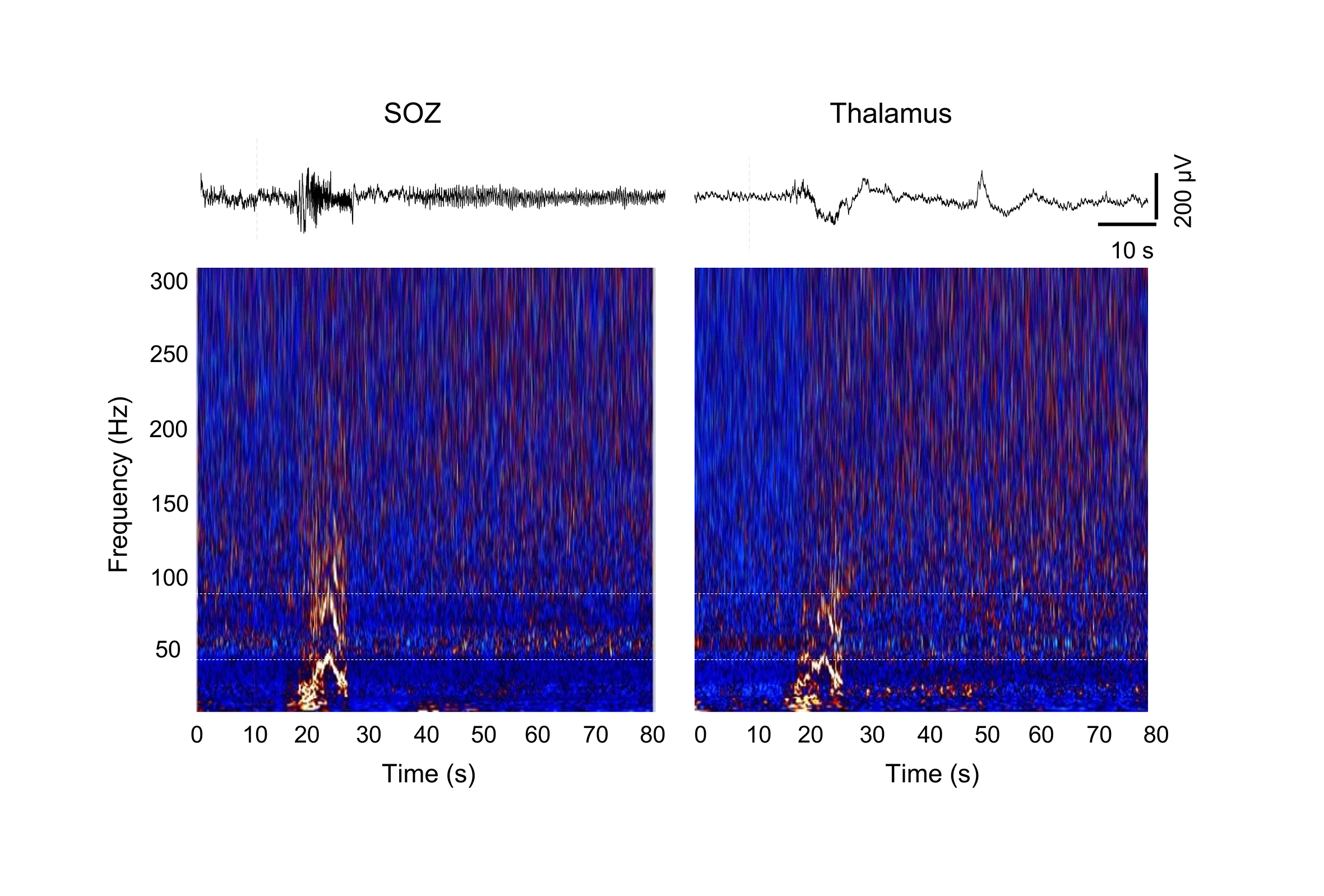

**Supplementary Figure 4 Illustration of EEG and corresponding H pattern for SOZ and thalamus in a patient.** The seizure originated from left medial frontal lobe. Simultaneous presence of H pattern is in the SOZ and ipsilateral thalamus, with the same frequency interval.

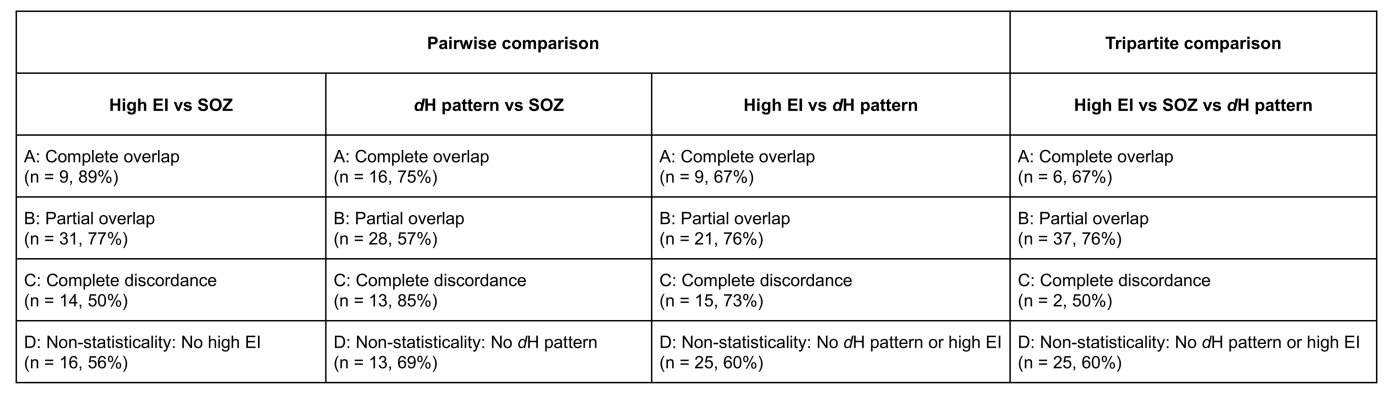

**Supplementary Figure 5: Concordance of high EI, SOZ, and *d*H pattern.** The concordance among these three markers was evaluated based on their EZq overlap and was categorized into four groups (A-D). The number of cases in each group and the proportion of patients with favorable outcomes were shown in parentheses.

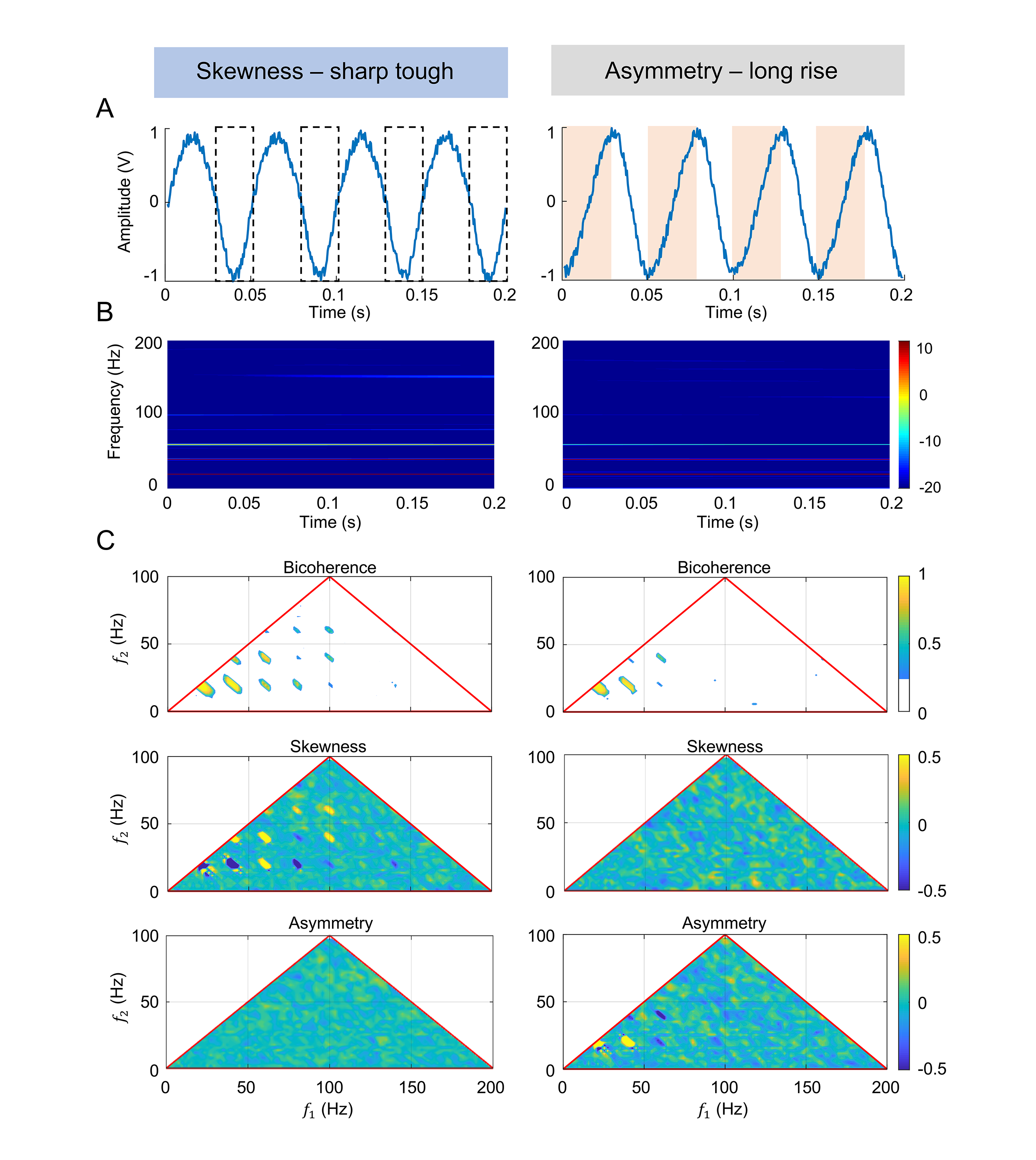

**Supplementary Figure 6 Two simulated waves to validate harmonic components induced by waveform distortion. (A)** Left: to illustrate the waveform that generates H pattern with sharp troughs, we constructed a sinusoidal wave with T_trough_ = 0.02 s and T_peak_= 0.03 s, resulting in a wave with a primary period T= 0.05 s. Right: to demonstrate the asymmetric waveform that generates H pattern, we constructed a sinusoidal wave with T_rise_ = 0.03 s and T_decay_ = 0.02 s, resulting in an asymmetric waveform with a primary period T= 0.05 s. **(B)** TFM for the two simulated waves. **(C)** Bispectral analysis for the two simulated waves. TFM: time frequency map.

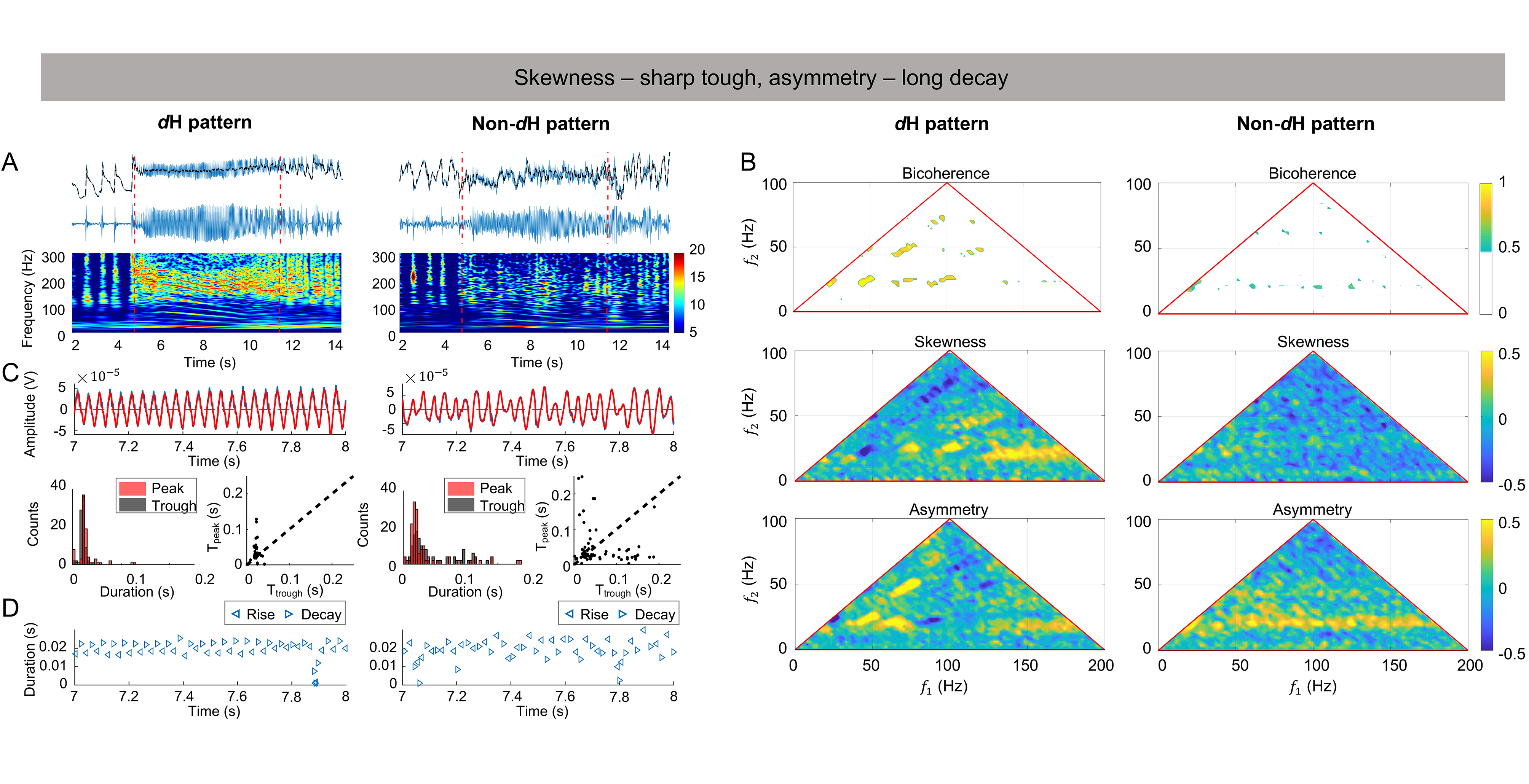

**Supplementary Figure 7 The *d*H pattern attributed to stronger skewness and asymmetry of the FA waves. (A)** Original SEEG signals in the *d*H/non-*d*H patterns (top), detrended SEEG signals (middle), TFM (bottom). **(B)** Bispectral analysis for the *d*H/non-*d*H patterns. **(C)** Comparison of peak and trough. Top: peaks and troughs of the FA waves (blue lines) in the *d*H/non-*d*H patterns are separately fitted by sin waves (red lines), respectively; Bottom: histogram (left) and scatter plot (right) depicting the distribution of T_peak_ vs. T_trough_ for the *d*H/non-*d*H patterns. **(D)** Comparison of rise (left triangles) and decay (right triangles).

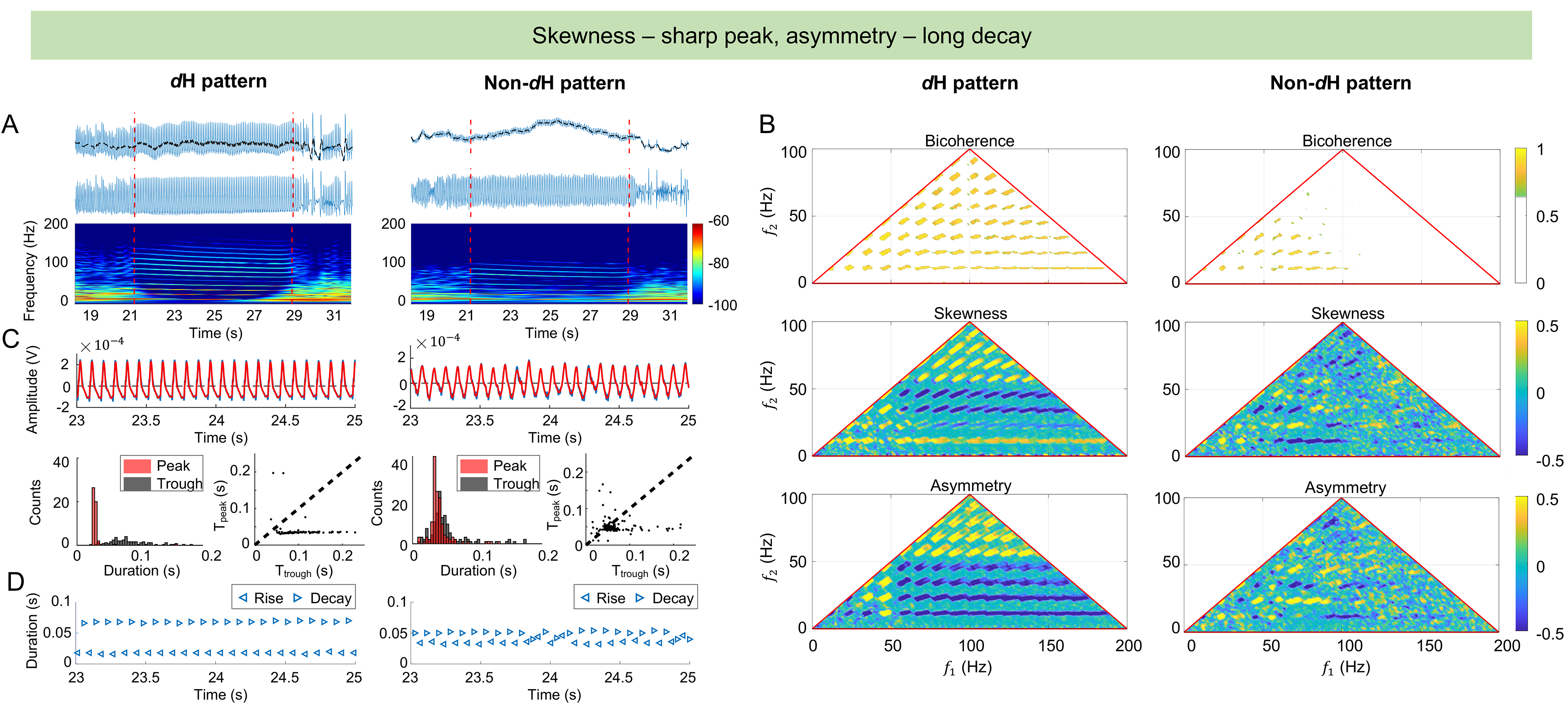

**Supplementary Figure 8 The *d*H pattern attributed to stronger skewness and asymmetry of the PS waves. (A)** Original SEEG signals in the *d*H/non-*d*H patterns (top), detrended SEEG signals (middle), and TFM (bottom). **(B)** Bispectral analysis for the *d*H/non-*d*H patterns. **(C)** Comparison of peak and trough. Top: peaks and troughs of the PS waves (blue lines) in the *d*H/non-*d*H patterns are separately fitted by sin waves (red lines), respectively; Bottom: histogram (left) and scatter plot (right) depicting the distribution of T_peak_ vs. T_trough_ for the *d*H/non-*d*H patterns. **(D)** Comparison of rise (left triangles) and decay (right triangles).

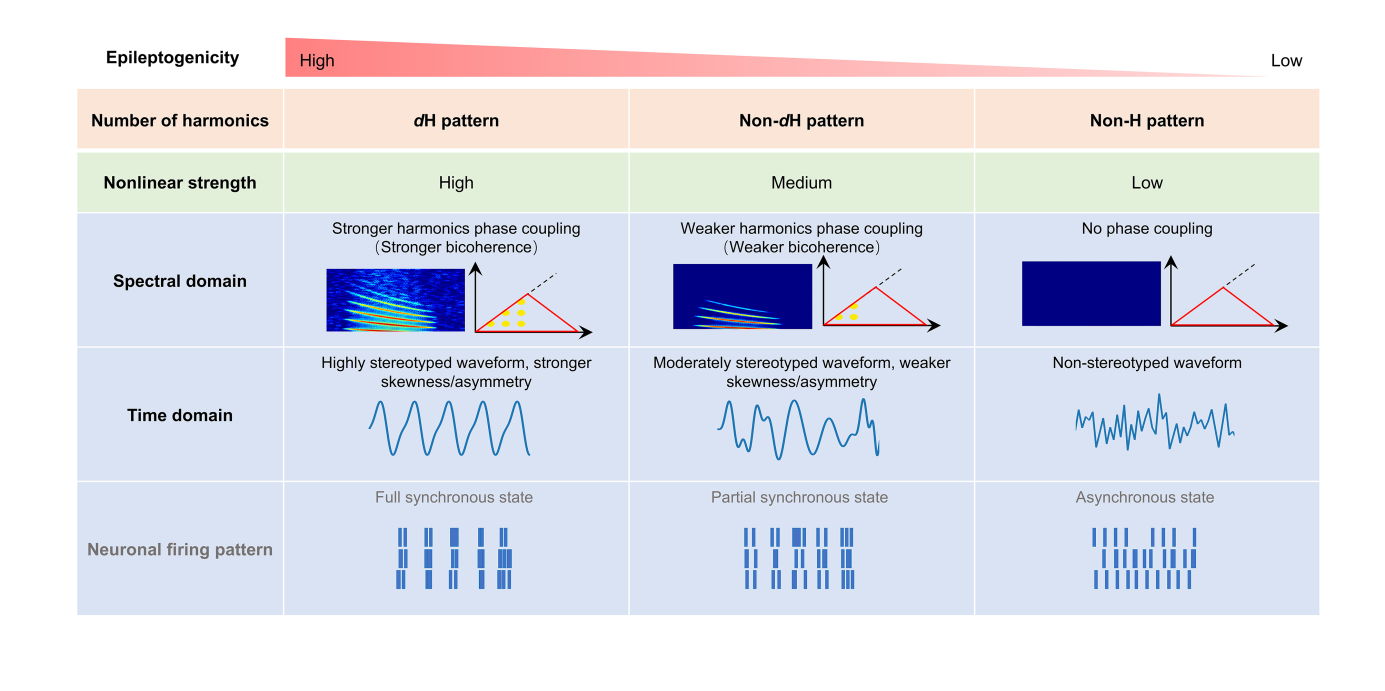
 **Supplementary Figure 9 Summary of the nonlinear features for H pattern.** The epileptogenicity, as reflected by the nonlinear features of H pattern, exhibits a clear gradient of decrease from the *d*H pattern to non-*d*H pattern, and is notably diminished in the non-H pattern. In the spectral domain, this is seen as a decline in harmonic numbers in TFM and coupled peaks in bispectral analysis. In the time domain, the decline is marked by reduced skewness and/or asymmetry of waveforms, with the *d*H pattern correlated with more stereotyped waveforms. It is hypothesized that the harmonic nonlinearity reflects the synchronization level of neuronal firing, which decreases from the *d*H pattern to non-*d*H pattern and is absent in the non-H pattern.
